# Inhaled booster vaccination with an MVA-based SARS-CoV-2 vaccine candidate induces T cell responses in lung

**DOI:** 10.1101/2025.10.09.25337642

**Authors:** Swantje I. Hammerschmidt, Rodrigo Gutierrez Jauregui, Joana Barros-Martins, Ivan Odak, Lennart Riemann, Verena Krähling, Anika Buchholz, Mahnaz Badpa, Georgia Kalodimou, Alina Tscherne, Michaela Friedrichsen, Inga Ravens, Jasmin Ristenpart, Anja Schimrock, Larissa Kasperek, Simon Schröder, Antonia Zapf, Christine Falk, Stephan Becker, Gerd Sutter, Asisa Volz, Christoph Schindler, Jens M. Hohlfeld, Reinhold Förster

**Affiliations:** Institute of Immunology, Hannover Medical School, Hannover, Germany; Department of Pathology, Molecular and Cell-Based Medicine, Icahn School of Medicine at Mount Sinai, New York, USA; The Tisch Cancer Institute, Icahn School of Medicine at Mount Sinai, New York, USA; Department of Pediatric Pneumology, Allergology and Neonatology, Hannover Medical School, Hannover; Philipps-Universität Marburg, Fachbereich Medizin, Institut für Virologie, Marburg, Germany; German Center for Infection Research (DZIF), Partner Site Gießen-Marburg-Langen, Marburg, Germany; Institute of Medical Biometry and Epidemiology, University Medical Center Hamburg-Eppendorf, Hamburg, Germany; Division of Virology, Department of Veterinary Sciences, LMU Munich, Oberschleißheim, Germany; German Centre for Infection Research (DZIF), Munich partner site, Munich, Germany; German Centre for Infection Research (DZIF), Translational Project Management Office, Braunschweig, Germany; Institute of Transplant Immunology, Hannover Medical School, Hannover, Germany; Institute of Virology, University of Veterinary Medicine Hannover, Hannover, Germany; German Centre for Infection Research (DZIF), Partner Site Braunschweig-Hannover, Hannover, Germany; Center for Clinical Trials (ZKS), Hannover Medical School, Hannover, Germany; Division of Airway Research, Fraunhofer Institute for Toxicology and Experimental Medicine ITEM, Hannover, Germany; Department of Respiratory Medicine and Infectious Disease, Hannover Medical School, Hannover; German Center for Lung Research (DZL), BREATH, Hannover, Germany; Cluster of Excellence RESIST (EXC 2155), Hannover Medical School, Hannover, Germany

**Author notes:** Corresponding author: Prof. Dr. Reinhold Förster Institute of Immunology Hannover Medical School Carl-Neuberg-Str. 1 D-30625 Hannover, Germany. Deceased author (Gerd Sutter). Trial registration: ClinicalTrials.gov NCT05226390. Funding: MWK Niedersachsen (14-76103-184 CORONA-11/20); DZIF (TTU 01.934 and FF 01.941); DFG (EXC 2155 “RESIST”); DZL (grant 82DZL002B1).

**Keywords:** inhalation, mucosal vaccines, SARS-CoV-2, COVID-19 vaccines, booster vaccination

## Abstract

**Background:** Parenteral COVID-19 vaccines induce strong systemic immunity, but they do not typically trigger pronounced respiratory immunity. In this context, mucosally applied vaccines might help to induce local immune responses for early viral clearance and reduced viral transmission.

**Methods:** In this investigator-initiated, open-label single-dose phase I trial, we analyzed the immunogenicity and safety of the vaccine candidate MVA-SARS-2-ST administered as an inhalation boost in COVID-19-immunized adults (n=23). MVA-SARS-2-ST represents a replication-deficient vector vaccine candidate built on the recombinant Modified Vaccinia virus Ankara (MVA) platform and expresses a prefusion-stabilized version of the full-length spike glycoprotein of SARS-CoV-2.

**Results:** While there was no increase in spike-specific antibodies in the blood, the inhalation of 10^7^ infectious units (IU) MVA-SARS-2-ST led to an increase in IFN-γ release after re-stimulation of whole blood with spike peptides. This enhanced IFN-γ release peaked at day 7 and remained detectable for at least 140 days after vaccination. Notably, selectively individuals with a history of COVID-19 (nucleocapsid protein (NCP)-seropositive study participants), but not individuals without a history of COVID-19 (NCP-seronegative study participants), showed a trend towards increased spike-specific IgA in the lung after inhalation of 10^7^ IU MVA-SARS-2-ST. In contrast, inhaled application of MVA-SARS-2-ST robustly induced spike-specific CD4+ and CD8+ T cell responses in the lung in both NCP-seronegative and NCP-seropositive individuals.

**Conclusions:** Collectively, our study demonstrated that a single booster inhalation of 10^7^ IU MVA-SARS-2-ST did not have a relevant impact on the humoral immune response, but induced specific T cell responses in blood and lung.

## Introduction

In response to the global impact of the SARS-CoV-2 outbreak in 2020, an unprecedented research effort was initiated, ultimately leading to the rapid development of several vaccines against the newly emerged coronavirus. The first SARS-CoV-2 vaccines to be developed were mRNA- and adenoviral vector-based vaccines as well as inactivated vaccines that were administered via the conventional intramuscular route and induced strong systemic immune responses (1–3). Through this means, these vaccines effectively mitigated the detrimental effects of the pandemic by reducing severe illness and mortality in COVID-19 (4, 5).

While injectable COVID-19 vaccines induce a strong systemic immune response, they do not typically trigger pronounced respiratory immunity (6, 7). However, since the respiratory tract is the primary entry site for SARS-CoV-2, mucosal IgA, as well as tissue-resident T and B cells, provide frontline immune defense by early viral clearance, inhibition of viral replication and reduction of onward transmission to others (8–10). This is particularly important for protecting immunocompromised and elderly individuals from severe disease. Furthermore, considering that viral spread can fuel the development of virus variants, reducing viral transmission can have pivotal public health implications especially during the emergence of a new epidemic pathogen. Therefore, mucosal vaccines have the potential to close the gap in the conventional intramuscular vaccination strategy by targeting effector cells and molecules to the mucosal site and thereby eliciting potent local immunity. Several already licensed mucosal vaccines show the promise of this application route (11).

In this context, we investigated the aerosol vaccination with an MVA-based COVID-19 vaccine candidate. MVA is a highly attenuated strain of vaccinia virus that efficiently infects, but cannot replicate in human cells. In its non-recombinant form, MVA is licensed as a vaccine against smallpox and Mpox. Furthermore, several recombinant MVA candidate vaccines have been tested successfully in clinical trials, with a recombinant MVA encoding four viral proteins of the virus family *Filoviridae* being approved in a heterologous prime-boost regimen against Ebola virus (12). The SARS-CoV-2 outbreak prompted the development of MVA-based vaccine candidates: among these, MVA-SARS-2-S encodes the full-length native SARS-CoV-2 spike protein (13), while MVA-SARS-2-ST encodes a pre-fusion-stabilized spike protein with an inactivated S1/S2 furin cleavage site (14). Phase I clinical trials evaluating the intramuscular administration of MVA-SARS-2-S and MVA-SARS-2-ST found no safety concerns and revealed that MVA-SARS-2-ST was more immunogenic than its predecessor MVA-SARS-2-S, although both being less immunogenic compared to licensed mRNA- and adenoviral-based vaccines in SARS-CoV-2 naïve individuals (15, 16).

Our preclinical studies in mice and hamsters demonstrated that intramuscular priming followed by respiratory tract boosting with MVA-SARS-2-S was a promising approach for the induction of local, respiratory as well as systemic immune responses capable of protecting against SARS-CoV-2 infections (17). These findings provided a solid foundation for this single-dose phase I trial, in which we evaluated the safety, reactogenicity, and immunogenicity of the COVID-19 vaccine candidate MVA-SARS-2-ST administered as an inhalation boost in SARS-CoV-2 immunized adults.

Our longitudinal analysis of the immune response revealed that the inhaled administration of 10^7^ IU MVA-SARS-2-ST induced a strong, spike-specific CD4 and CD8 T cell response in blood and lung tissues. However, this approach appears to be not suited in inducing binding or neutralizing antibodies.

## Results

### Inhalation of 10^7^ IU MVA-SARS-2-ST did not induce spike-specific antibody responses in blood

In this single-dose phase I trial, we analyzed the safety, reactogenicity, and immunogenicity of the vaccine candidate MVA-SARS-2-ST administered as inhalation boost in SARS-CoV-2 immunized adults. As reported elsewhere, we found that a single booster inhalation of 10^7^ IU MVA-SARS-2-ST had an acceptable safety and tolerability profile in healthy volunteers (n=23) (18).

The secondary objective of this phase I trial was to assess spike-specific antibody responses in blood. To this aim, we longitudinally measured anti-S1-IgG in serum samples by ELISA. As reported earlier, all participants had detectable S1-specific IgG before inhalation (18), in accordance with their vaccination history and the inclusion criterion to show S1-specific IgG titers between 10 and 1200 RU/mL (corresponding to 32 – 3840 BAU/mL) prior to inhalation. While there was no increase in S1-binding IgG during the first 14 days after application, some participants showed a strong increase of anti-S1-IgG between 56 and 140 days after inhalation (18). Presumably, this increase was not related to the inhalation of the vaccine candidate MVA-SARS-2-ST, but may have resulted from a SARS-CoV-2 breakthrough infection. To explore this possibility, we tested the serum samples for anti-NCP-IgG. MVA-SARS-2-ST does not encode the nucleocapsid protein (NCP). Thus, monitoring the levels of anti-NCP-IgG provides the opportunity to detect SARS-CoV-2 breakthrough infections, which have occurred before or during the phase I study. Based on the levels and the dynamics of NCP-specific IgG, we assigned the study participants to three different groups to make the immunogenicity analysis more interpretable (Figure 1A). Many study participants (n=11, color-coded in orange) were NCP-seronegative at the start of the study and stayed NCP-seronegative throughout the whole study (Figure 1A). These participants did not have a SARS-CoV-2 infection before the study and most likely did not experience a breakthrough infection after the inhalation boost. Other study participants (n=4, color-coded in green) were initially NCP-seronegative at the start of the phase I study, but turned NCP-seropositive between 56 and 140 days after inhalation (Figure 1A). One of these study participants showed only a moderate increase in anti-NCP-IgG, but this coincided with a strong increase in S1-specific IgG most likely related to SARS-CoV-2 infection (Figure 1B). In line with this, three of these study participants, including the individual with only moderate increase in anti-NCP-IgG, reported a breakthrough infection at study day 65, 97 or 99, respectively (18). Thus, we concluded that these participants did not have a SARS-CoV-2 infection before the study, but experienced a breakthrough infection after the inhalation boost. Finally, other study participants (n=8, color-coded in purple) were NCP-seropositive already at the beginning of the study (Figure 1A), indicative for SARS-CoV-2 infection(s) in the past. The high levels of NCP-specific IgG at the start of the study made it difficult to detect any meaningful changes throughout the study, which could indicate a breakthrough infection. Thus, based on the anti-NCP-IgG levels, we could not further distinguish in this group between participants with or without a breakthrough infection after the inhalation boost. However, it should be noted that none of these participants reported a breakthrough infection throughout the study.

**Figure 1:**
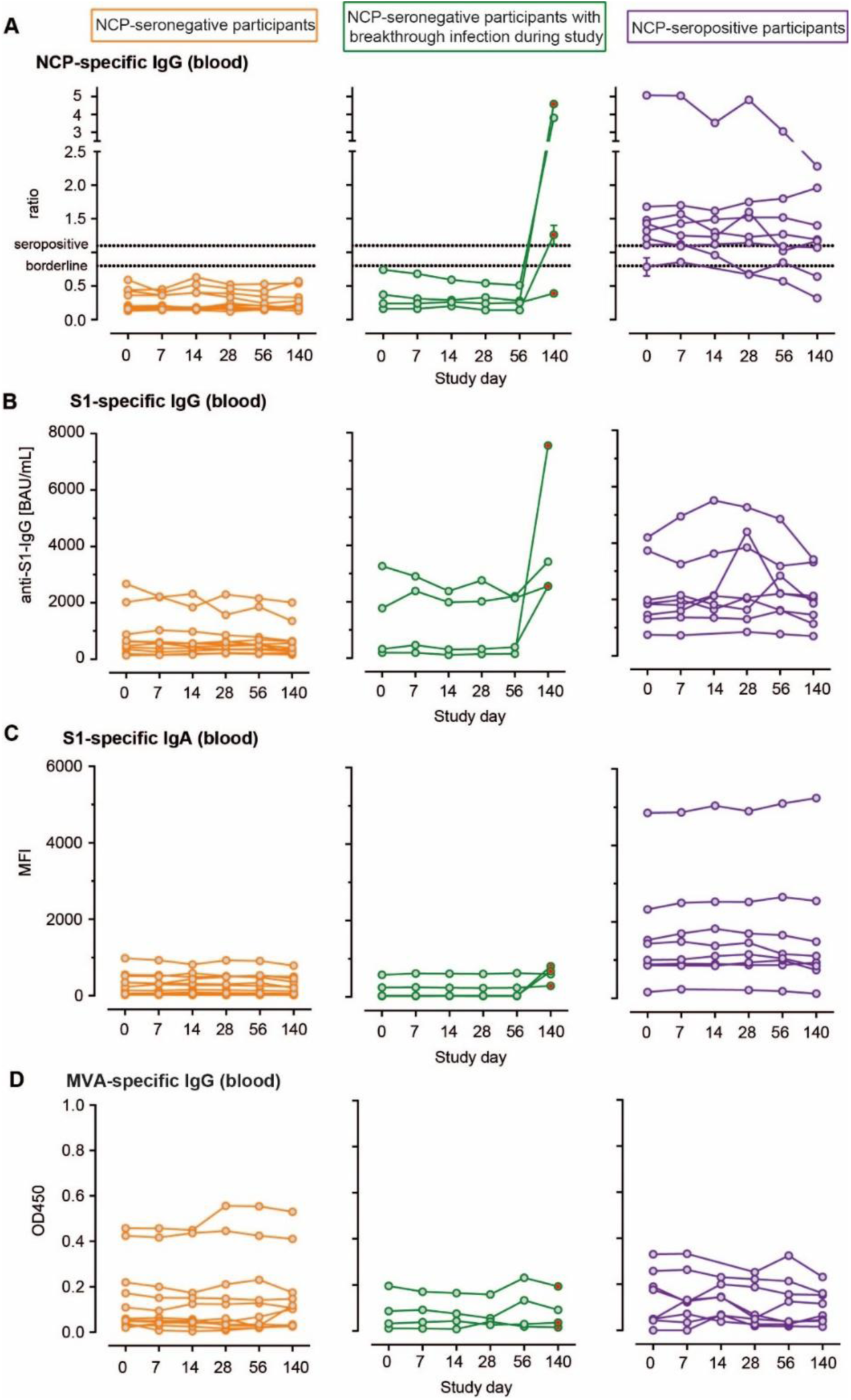
Analysis of the humoral response in blood after MVA-SARS-2-ST inhalation. Anti-NCP-IgG ratios (A), anti-S1-IgG (B), anti-S1-IgA (C) and anti-MVA-IgG levels (D) measured between day 0 and day 140 in the serum of participants without SARS-CoV-2 breakthrough infection (orange circles), participants with breakthrough infection during the study (green circles), and participants with previous breakthrough infection before the start of the study (purple circles). Red-filled circles represent samples after reported breakthrough infection. BAU, binding antibody units; MFI, mean fluorescence intensity; OD450, optical density measured at 450 nm. Summary statistics are provided in Supplementary Tables S1 through S6.

Upon dividing the study participants into the three groups described above, it became evident that at the outset of the study, the vaccinated participants with prior SARS-CoV-2 infection exhibited higher levels of S1-binding IgG and IgA antibodies compared to the vaccinated participants without prior SARS-CoV-2 infection (Figure 1B, C). Neither of the groups showed dynamic changes in S1-specific IgG or IgA early after inhalation (Figure 1B, C). Consistently, serum samples did not show an increase in neutralizing activity against SARS-CoV-2 during the first 28 days after inhalation (Figure S1). Furthermore, the increase in S1-binding IgG and IgA levels between day 56 and 140 in some of the study participants was found to coincide with an increase of anti-NCP-IgG (Figure 1 A, B), suggesting that breakthrough infections were a key driver of anti-S1-IgG induction.

Collectively, our data suggest that inhalation of 10^7^ IU MVA-SARS-2-ST did not have a relevant impact on the humoral response in blood. Notably, we were unable to detect anti-vector immunity, as there was no induction of MVA-specific IgG (Figure 1D).

### Inhalation of MVA-SARS-2-ST induced a systemic cellular immune response detectable as increased IFN-γ release after re-stimulation of whole blood

In addition to the humoral response, we evaluated the cellular immune response before and after vaccine inhalation. First, we conducted an IFN-γ release assay (IGRA) to measure the secretion of IFN-γ by spike-specific T cells. Our results showed that inhalation of MVA-SARS-2-ST led to an increase in IFN-γ release after re-stimulation of whole blood with spike peptides (Figure 2A). NCP-seronegative and NCP-seropositive study participants exhibited a similar response (Figure 2A). The enhancement of IFN-γ release peaked at day 7 and gradually declined until day 140 (Figure 2A). Notably, the levels of re-stimulation-triggered IFN-γ at 14 days post MVA-SARS-2-ST inhalation (median 3382 mIU/mL; IQR 6336) were comparable to those at 14 days post intramuscular boosting with the BNT162b2 mRNA vaccine (median 3222 mIU/mL; IQR 6898) (19). In contrast to enhanced IFN-γ release, whole blood re-stimulation did not result in enhanced TNF-α secretion post inhalation as shown in Figure S2A. Subsequently, we analyzed the frequencies of spike-specific T cells in blood using flow cytometry. Following PBMC stimulation with spike peptides, we did not observe an expansion of IFN-γ and/or TNF-α producing CD4+ or CD8+ T cells post vaccination (Figure 2B).

**Figure 2:**
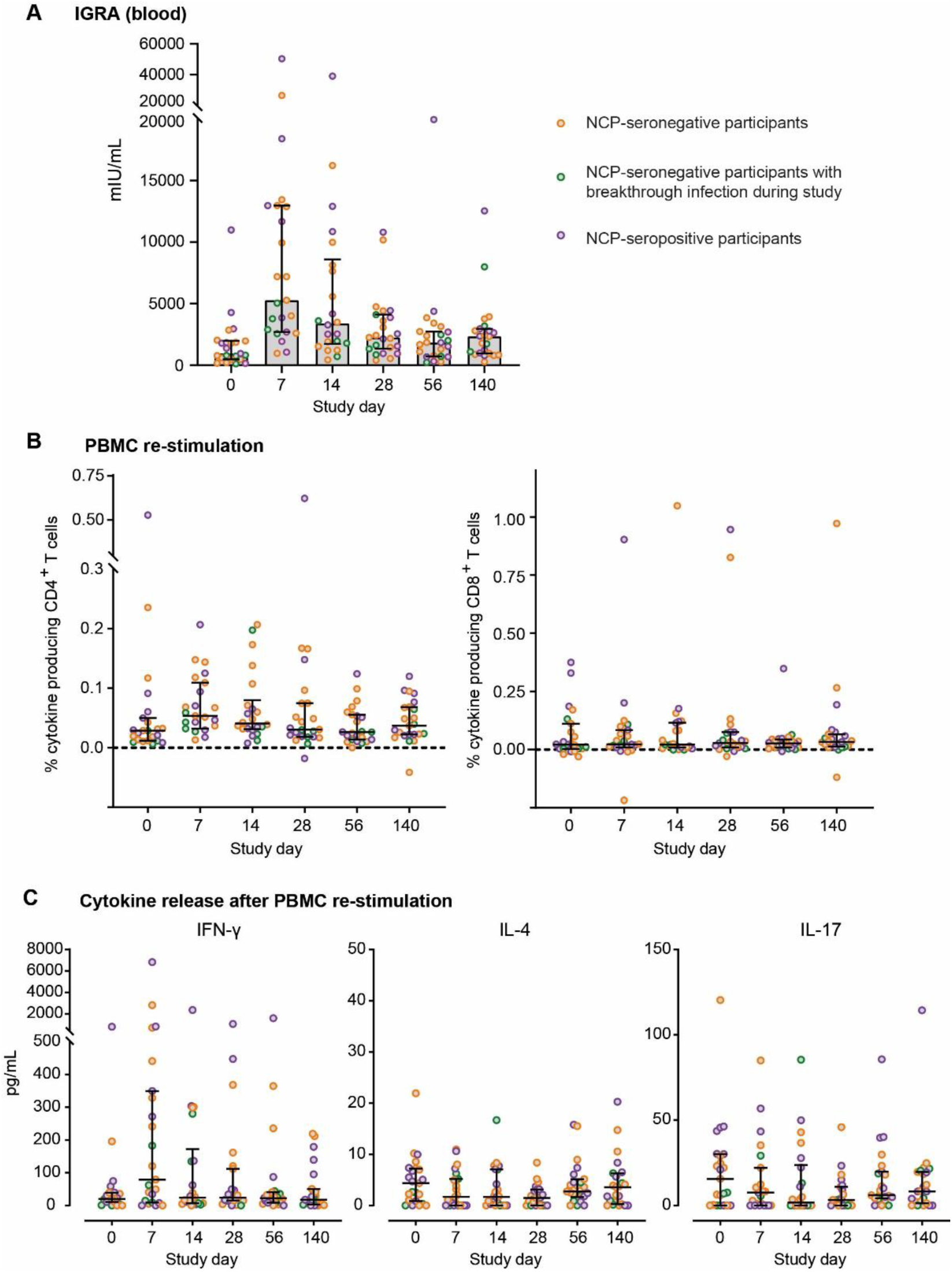
Analysis of the T cell response in blood after MVA-SARS-2-ST inhalation. (A) Concentrations of IFN-γ in whole blood supernatants after stimulation with SARS-CoV-2 S1 domain for 20–24h measured by IGRA (Euroimmun). (B) Percentage of cytokine-secreting CD4+ and CD8+ T cells after PBMC stimulaton with Spike peptide. We calculated the total number of cytokine-secreting cells as the sum of IFNγ+ TNFα−, IFNγ+ TNFα+ and IFNγ− TNFα+ cells. For each sample, the background signal of the DMSO condition was deducted from the signal of the Spike stimulation. (C) Cytokine release after re-stimulation of peripheral blood mononuclear cells (PBMC) with spike peptides. (A-C) Lines represent group median with interquartile range; dots represent individual participants: participants without SARS-CoV-2 breakthrough infection (orange circles), participants with breakthrough infection during the study (green circles), and participants with previous breakthrough infection before the start of the study (purple circles). Summary statistics are provided in Supplementary Tables S7 through S12.

However, when profiling the immune response of spike-re-stimulated PBMCs by quantifying the released cytokines in the supernatants, we observed increased concentrations of IFN-γ 7 days post inhalation (Figure 2C). The levels of the cytokines IL-4 and IL-17 remained consistently low (Figure 2C). Notably, there was no increase in IL-2, IL-10, IL-12p70 and or TNF-α secretion after PBMC re-stimulation (Figure S2B). Interestingly, levels of CXCL9 and CXCL10 appear to increase upon stimulation early post inhalation, which may be in response to IFN-γ secretion (Figure S2B).

Collectively, our data suggest that inhalation of 10^7^ IU MVA-SARS-2-ST induced a systemic cellular immune response characterized by increased IFN-γ release after re-stimulation of whole blood. This enhanced IFN-γ secretion peaked at day 7 and remained detectable until the end of the study, i.e. up to 140 days post vaccination. Furthermore, stimulated PBMC exhibited a Th1 cytokine profile with enhanced secretion of IFN-γ at day 7 and marginal release of the Th2-cytokine IL-4 and the Th17-cytokine IL-17.

### Inhaled application of MVA-SARS-2-ST slightly increased spike-specific IgA exclusively in the lungs of NCP-seropositive individuals, but not in the lungs of NCP-seronegative individuals

The second main immunological objective of the study was to assess anti-S1 antibody responses in BAL fluids. In ELISA, we were unable to detect any changes in S1-binding IgG in BAL fluids (Figure 3A). Next, we analyzed the levels of S1-binding IgA in BAL and found that S1-specific IgA was predominantly present in the BAL of study participants who had experienced a SARS-CoV-2 infection before enrollment into the trial, while the majority of SARS-CoV-2 infection-naïve study participants lacked detectable amounts of spike-specific IgA, despite being vaccinated with licensed vaccines three times (Figure 3B). Interestingly, inhalation of MVA-SARS-2-ST slightly increased mucosal spike-specific IgA selectively in NCP-seropositive study participants, while the NCP-seronegative study participants remained low in pulmonary IgA (Figure 3B). Notably, induction of spike-specific IgA in the BAL did not coincide with anti-MVA-IgA induction in the BAL (Figure 3C). In the large majority of BAL samples, we were unable to detect any neutralizing activity (data not shown).

**Figure 3:**
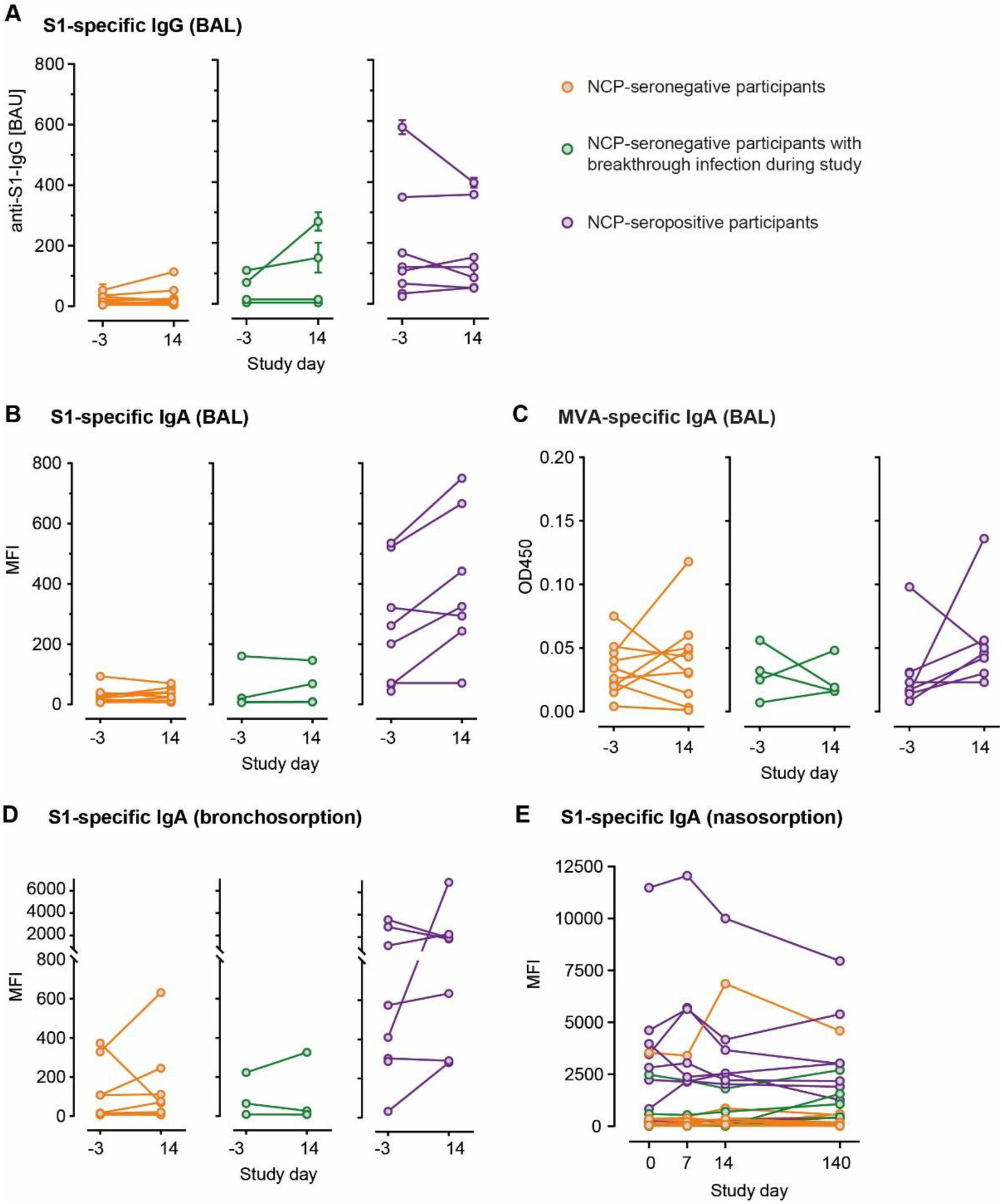
Analysis of the mucosal humoral response after MVA-SARS-2-ST inhalation. Anti-S1-IgG (A), anti-S1-IgA (B), and anti-MVA-IgG levels (C) measured before and 14 days post inhalation in the bronchoalveolar lavage (BAL). Anti-S1-IgA measured before and after inhalation in bronchosorption (D) and nasosorption (E). Participants without SARS-CoV-2 breakthrough infection (orange circles), participants with breakthrough infection during the study (green circles), and participants with previous breakthrough infection before the start of the study (purple circles). MFI, mean fluorescence intensity; OD450, optical density measured at 450 nm. Summary statistics are provided in Supplementary Tables S13 through S24.

Beyond BAL, we also conducted bronchosorption and nasosorption sampling. Bronchosorptions confirmed the presence of elevated levels of S1-specific IgA in NCP-seropositive individuals (Figure 3D and E). However, unlike BAL, we did not detect any immunologically relevant changes in the levels of spike-specific IgA in bronchosorption or nasosorption samples (Figure 3D and E). This may be due to the very small and confined sampling area of the bronchosorption and nasosorption filters.

Together, selectively NCP-seropositive study participants, but not NCP-seronegative individuals, show a trend towards increased spike-specific IgA in the lung after inhalation of 10^7^ IU MVA-SARS-2-ST.

### Aerosol vaccination with MVA-SARS-2-ST induced spike-specific CD4^+^ and CD8^+^ T cells in the lung

Given the rationale behind the inhaled application which aims to target the mucosal immune system, we assessed the T cell response in the lung. To this end, we isolated cells from the BAL and re-stimulated the cells with spike peptides in the presence of brefeldin A. Intracellular flow cytometry staining revealed that the frequencies of IFN-γ and/or TNF-α producing CD4^+^ as well as CD8^+^ T cells in BAL samples expanded 14 days post inhalation (Figure 4A). Upon spike stimulation in the absence of Brefeldin A, BAL cells of some study participants released strongly increased levels of IFN-γ, while the cells of other study participants showed a weak to moderate IFN-γ response 14 days post inhalation (Figure 4B). For IL-4 and IL-17, the study participant population responded more uniformly with some minor increase after inhalation (Figure 4B). Despite the increase, IL-4 and IL-17 remained in a low concentration range after inhalation. Similarly, IL-10 and IL-12p70 levels increased within a low range (Figure S3). IL-2 secretion followed a similar pattern like IFN-γ with a broad spectrum spanning strong to week re-stimulation-triggered IL-2 responses post inhalation (Figure S3).

**Figure 4:**
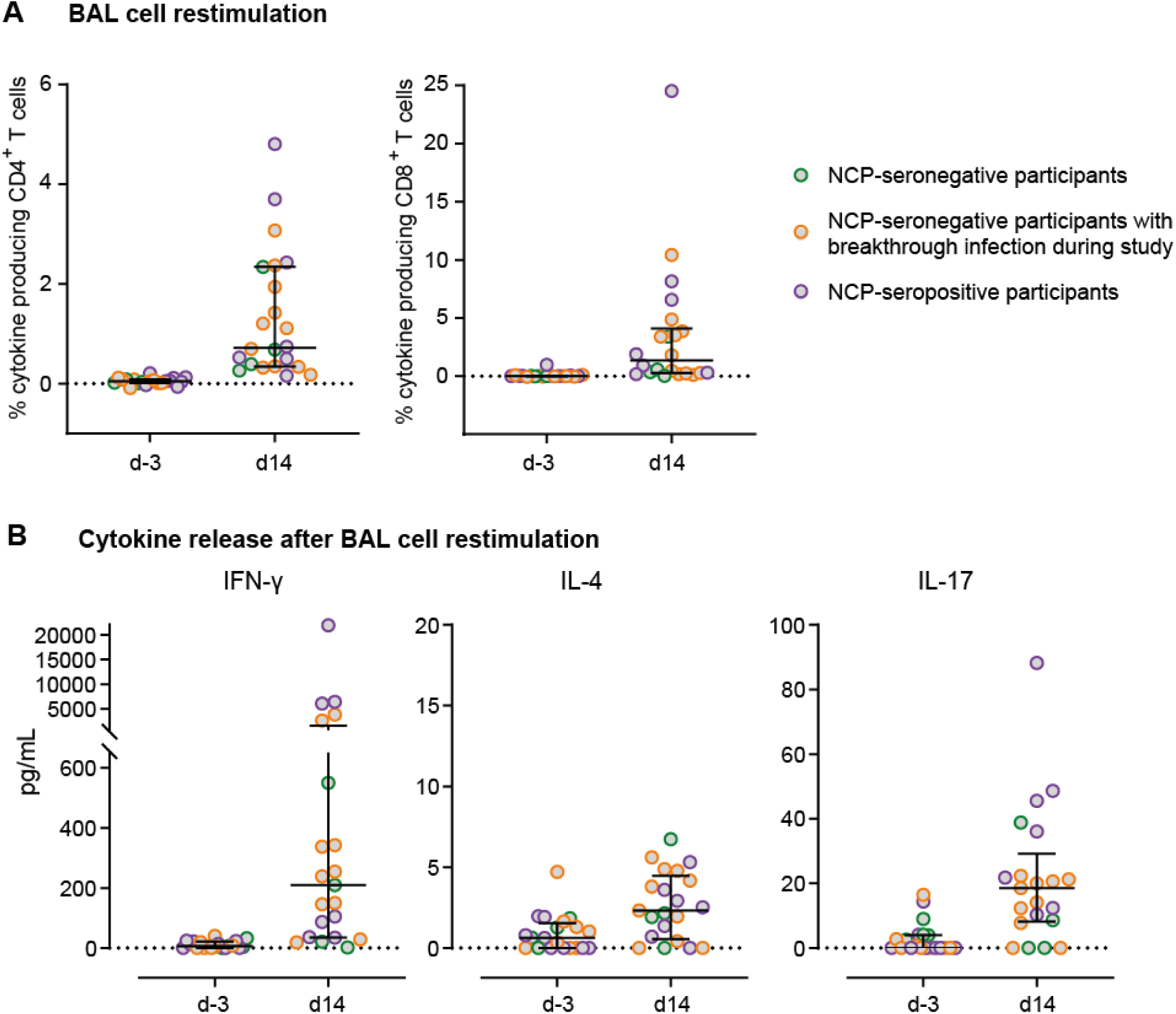
Analysis of the T cell response in lung after MVA-SARS-2-ST inhalation. (A) Percentage of cytokine-secreting CD4+ and CD8+ T cells after BAL cell stimulation with Spike peptide. We calculated the total number of cytokine-secreting cells as the sum of IFNγ+ TNFα−, IFNγ+ TNFα+ and IFNγ− TNFα+ cells. For each sample, the background signal of the DMSO condition was deducted from the signal of the Spike stimulation. (B) Cytokine release after re-stimulation of BAL cells with spike peptides. (A-B) Lines represent group median with interquartile range; dots represent individual particpants: participants without SARS-CoV-2 breakthrough infection (orange circles), participants with breakthrough infection during the study (green circles), and participants with previous breakthrough infection before the start of the study (purple circles). Summary statistics are provided in Supplementary Tables S25 through S29.

Notably, no increase in TNF-α secretion was detected after PBMC re-stimulation (Figure S3) suggesting that the increase of spike-specific cytokine-producing T cells predominantly relied on the increase of IFN-γ secreting rather than TNF-α secreting T cells. Furthermore, levels of CXCL9 and CXCL10 increased upon stimulation post inhalation, possibly in response to IFN-γ secretion (Figure S3).

Collectively, our data suggest that inhalation of 10^7^ IU MVA-SARS-2-ST induced spike-specific CD4^+^ and CD8^+^ T cell responses in the lung.

## Discussion

In this phase I clinical trial, we evaluated the safety and immunogenicity of the vaccine candidate MVA-SARS-2-ST administered as an inhalation boost in SARS-CoV-2 immunized adults. Our results showed that a single booster inhalation of 10^7^ IU MVA-SARS-2-ST had an acceptable safety and tolerability profile in healthy volunteers (n=23), as reported in detail by Hohlfeld at al. (18). No serious adverse events were observed and no abnormalities were detected in safety laboratory measurements (18). Pulmonary function analysis revealed a small, temporary and spontaneously reversible decline on day 14 (18).

In addition to evaluating the safety and tolerability as primary objectives, we also assessed the immunogenicity and monitored humoral and cellular responses in blood and lung. Notably, inhalation of 10^7^ IU MVA-SARS-2-ST did not induce spike-specific antibodies in blood which is in contrast to our preclinical studies in mice and hamsters where we readily observed the induction of neutralizing antibodies in serum and BAL samples after intramuscular priming followed by intranasal boosting (17). This divergent outcome may be due to species-related differences, as well as the different routes of respiratory tract boosting used in the preclinical studies (intranasal application) versus the clinical study (inhaled application). Furthermore, it is also possible that the lower dose/body weight ratio for humans compared to mice and hamsters may have contributed to the weaker induction of spike-specific antibody responses. When administered intramuscularly, MVA-SARS-2-ST readily induced S1-specific antibodies (16), suggesting that the weak humoral response post inhalation may depend on the administration route or the low dose per body weight ratio. Notably, we can exclude the generation of vector-specific antibodies as the underlying cause of weak induction of spike-specific antibody responses, as we could not detect induction of MVA-specific antibodies in blood or BAL samples.

While there was no vaccine-induced increase in spike-specific binding or neutralizing antibodies in blood, inhaled vaccination led to an increase in IFN-γ release after spike peptide re-stimulation of whole blood. This enhanced IFN-γ release peaked at day 7 and remained detectable at least up to 140 days. Similarly, after re-stimulation of peripheral blood mononuclear cells, there was an increase in the concentration of IFN-γ at day 7. Notably, the TH2-cytokine IL-4 and the TH17-cytokine IL-17 were only marginally detected in the supernatants of spike-stimulated PBMC, indicating a predominantly TH1-type immune response in the blood.

Given the rationale behind the inhaled application, which aims to target the mucosal immune system, we also analyzed the immune response in the lung. Regarding the mucosal antibody response, we detected a slight induction of spike-specific IgA, exclusively in the lungs of study participants previously infected with SARS-CoV-2. Notably, we detected no or only low neutralizing activity in the BAL fluids before or after inhalation, even in the study participants with prior breakthrough infection (data not shown). This limitation might be based on the large volume of 200 mL, which the BAL was performed with, and which strongly diluted the epithelial lining fluid, thereby hampering the detection of potentially present neutralizing antibodies. Nevertheless, similarly to the absent humoral response in blood, inhalation of 10^7^ IU MVA-SARS-2-ST may not have a strong effect on the humoral response in BAL, possibly due to the low dose. Regarding the induction of mucosal IgA, it is of interest that study participants with prior breakthrough infection responded better than infection-naïve individuals. SARS-CoV-2 infections have been reported to shape the immune landscape in the lung by inducing tissue-resident memory B cells (20), which might be readily activated by the inhaled vaccine leading to a more robust mucosal IgA response. However, for an intramuscular prime-intranasal boost regimen, it has been reported in mice that IgA^+^ B cells primarily arise through direct class switching from pre-existing IgG^+^ B cells in draining lymph nodes (21). This suggests that NCP-seronegative study participants should, in principle, be responsive to a mucosal boost, as they possess the necessary pre-existing IgG^+^ B cells that can undergo class switching to produce IgA^+^ B cells.

Intriguingly, inhaled vaccination with 10^7^ IU MVA-SARS-2-ST triggered T cell responses in the lung. Notably, in contrast to the pulmonary antibody response, NCP-seronegative and NCP-seropositive study participants responded equally well with regard to the mucosal T cell response. Similarly, a tuberculosis vaccine trial has shown that aerosol vaccination with MVA expressing a *M. tuberculosis* antigen induced mycobacteria-specific mucosal and systemic cellular immune responses (22). Given the challenges posed by antibody evasion by rapidly evolving pathogens, the induction of mucosal T cell memory has emerged as a critical objective for the development of next-generation vaccines. Mucosal T cell memory has several advantages. For example, compared to blood, spike-reactive tissue memory T cells persist for longer periods of times post-vaccination (23) and provide faster viral clearance (7). Several studies have provided evidence that intramuscular vaccination does not establish pulmonary T cell memory (6, 7). Therefore, inhaled application could potentially close this gap as part of a ‘‘prime-pull’’ vaccination strategy which involves first eliciting systemic T cell responses through intramuscular priming and then recruiting T cells to the lung by respiratory delivery of vaccine boosters.

Along this line, a recent mouse study has shed light on the immune mechanisms that enable the conversion of a systemic immune response after intramuscular immunization to mucosal responses after intranasal boosting (24). The study demonstrated that CD4^+^ T cells in the lung induced the expression of the chemokines CXCL9 and CXCL10, which is crucial for the recruitment of memory B cells to the lung (24). Notably, we also observed the presence of CXCL9 and CXCL10 in the supernatant of re-stimulated BAL cells post inhalation. This suggests that the inhaled application of 10^7^ IU MVA-SARS-2-ST may have set the stage for faster and enhanced B cell recruitment in the event of subsequent breakthrough infections.

Collectively, inhalation of 10^7^ IU MVA-SARS-2-ST did not induce an increase in binding or neutralizing spike-specific IgG antibodies in blood, but generated a long-lasting T cell response in blood as detected by increased IFN-γ release after spike-specific re-stimulation of whole blood up to 140 days after inhalation. Notably, inhalation of 10^7^ IU MVA-SARS-2-ST slightly increased spike-specific IgA exclusively in the lungs of NCP-seropositive study participants, but not in the lungs of NCP-seronegative study participants. Furthermore, inhalation of 10^7^ IU MVA-SARS-2-ST robustly triggered specific CD4^+^ and CD8^+^ T cell responses in the lung of both NCP-seronegative as well as NCP-seropositive individuals. Spike-specific T cells in both compartments, blood and lung, exhibited a favorable cytokine profile with high IFN-γ release and marginal IL-4 and IL-17 secretion.

While there was no or only weak induction of antibodies in this trial, the application of MVA-based vaccine candidates may be a promising approach for scenarios where strong T cell responses in the lung are required, such as the treatment of tuberculosis, cancer, or other therapeutic vaccinations. Technically, the inhaled application could potentially be performed using mobile nebulizers that are widely available and relatively inexpensive.

This highly experimental clinical study has provided a unique opportunity to gain a deeper understanding of the immunogenicity of a vaccine candidate applied via inhalation. Throughout the course of the study, additional analyses such as high dimensional spectral flow cytometry, bulk mRNA sequencing, and 48-plex cytokine measurements were conducted. Upcoming systems biology analysis of these data will potentially reveal molecular signatures associated with higher immune responsiveness towards inhalation. Furthermore, comparison with intramuscular delivery of MVA-SARS-2-ST may reveal inhalation-specific immune mechanisms. Together, this will contribute to the development of future vaccination strategies that employ the respiratory route, which is of high relevance for the combat against future emerging respiratory pathogens.

## Methods

### Sex as a biological variable

Male and female participants were enrolled in this study, which was open to all sexes. 18 out of 23 (78.3%) participants were female. Sex was not considered as a biological variable.

### Study approval

The study protocol was reviewed and approved by the National Authority (Paul-Ehrlich-Institut, Langen, Germany) and the Ethics Committee of Hannover Medical School (EK 10012_AMG_mono_2021). The study was performed in compliance with the German Drug Law (AMG), German Good Clinical Practices (GCP) ordinance (GCP-V), ICH GCP, and the applicable national and European regulations covering the conduct of clinical studies. A local safety monitoring board provided clinical oversight. All participants provided written, informed consent. This trial was registered at ClinicalTrials.gov, NCT05226390, and EudraCT, 2020-004010-35.

### Study design

This was an investigator-initiated, single-center, open-label phase I trial to evaluate the safety, reactogenicity and immunogenicity of the MVA-SARS-2-ST vaccine candidate administered as inhalation boost in SARS-CoV-2 immunized adults (1 x 10^7^ IU/dose MVA-SARS-2-ST in 0.5 mL as inhalation). We included men and women aged 18–60 years with no clinically significant health problems as determined during medical history and physical examination, a body-mass index of 18·5–30·0 kg/m² and weight of more than 50 kg at screening, normal pulmonary function (FEV1 predicted ≥ 80% and FEV1/FVC > 70%) and a negative pregnancy test for women. Further, participants had to be either immunized twice with any regimen using any EU-marketed SARS-CoV-2 vaccine (mRNA-, viral vector-, protein-based, or live attenuated SARS-CoV-2 vaccine) or with a single application of the COVID-19 Vaccine Janssen and optionally subsequently booster-immunized with any EU-marketed mRNA vaccine at least 3 months prior to enrollment. Participants were required to have SARS-CoV-2 specific IgG concentration between 10 RU/mL and 1200 RU/mL determined by Anti-SARS-CoV-2-QuantiVac-ELISA (IgG). Exclusion criteria contained current smoking/vaping or smoking/vaping in the previous year. The full list of inclusion and exclusion criteria are provided in (18). The trial was conducted at Hannover Medical School, Germany, from June 2022 to November.

### Procedures

The trial included a screening period from day -28 to day -1, a baseline period from day -7 to day -2, a single dose treatment on day 0 and a follow-up period until day 140. On day 0, participants received a single booster dose of 1 x 10^7^ IU MVA-SARS-2-ST in 0.5 mL as inhalation. Safety assessments, including pulmonary function testing, laboratory investigations, and vital signs, were conducted throughout the trial. Blood and nasosorption samples were collected throughout the trial. Two bronchoscopies with sampling of bronchoalveolar lavage (BAL) and bronchosorption were conducted at baseline and at day 14 post inhalation. The full schedule of assessments is published in (18). Bronchoscopic procedures adhered to published methods (25). To sample lining fluid, a bronchosorption catheter (Bronchosorption™ FX-I, Hunt Developments) was inserted through the working channel of the bronchoscope and placed onto the bronchial mucosa for 30 sec. After removing the bronchosorption catheter, the catheter tip was cut off and transferred into a Costar Spin-X vial (Corning) filled with 100 µL PBS/0.5% BSA. BAL was performed in a segment of the right middle lobe or lingula lobe using 200 mL pre-warmed sterile saline (0.9%) in 50 mL-aliquots. To sample lining fluid from the nasal cavity, Nasosorption™ FX-i devices (Hunt Developments) were placed into both nostrils for 2 min and subsequently transferred into tubes filled with 500 µL PBS/0.5% BSA. Lining fluid was extracted by vortexing and centrifugation in a Spin-X vial (Corning). Blood was collected using EDTA S-Monovette (Sarstedt). After plasma isolation, PBMCs were harvested from EDTA blood by Ficoll gradient centrifugation. Serum was collected using Gel monovettes (Sarstedt) and stored at −80 °C. Additionally, whole blood was collected in lithium heparin monovettes (Sarstedt) and used freshly for the interferon-gamma release assay.

### Vaccine candidate and administration via inhalation

MVA-SARS-2-ST is a vaccine candidate based on a recombinant MVA encoding a pre-fusion-stabilized spike protein with an inactivated S1/S2 furin cleavage site (14). The vaccine candidate was manufactured in DF-1 cells by IDT Biologika (Dessau, Germany). One vial with 1 x 10^7^ IU/dose MVA-SARS-2-ST in 0.5 mL per study participant was administered via inhalation. For this, the content of one vial was filled into a vibrating mesh nebulizer (M-neb dose+, NEBU-TEC med. Produkte Eike Kern GmbH). The study participants wore a nose-clip to avoid nasal breathing. The study participants inhaled the breath-triggered aerosol via a mouth piece. After a breath-hold of approximately 5 seconds, the study participants exhaled via a separate, filter-equipped mouth port to avoid release into the environment. Under observation and instruction of qualified staff, the subjects performed single inhalation maneuvers from the nebulizer with breath-actuation which released approximately 7 µL of liquid per inhalation. One dose of 0.5 mL MVA-SARS-2-ST was inhaled in around 70 inhalations within approximately 10 to 15 minutes.

### Enzyme-linked immunosorbent assays (ELISAs)

S1-binding IgG was measured as described previously (26). In detail, ELISA plates (Greiner bio-one) were coated with 1 μg/mL SARS-CoV-2 spike (S1) protein (#40591-V08H, Sino Biological) overnight at 4°C. After blocking with PBS + 5% milk powder, human sera were diluted 1:101 in PBST containing 1% milk powder and allowed to react with the S1 protein for 1 h. Polyclonal rabbit anti-human IgG HRP (#P0214, Agilent Technologies) was added to the plates and incubated for 30 min at room temperature (RT). 3,3′,5,5′-tetramethylbenzidine (TMB) substrate solution (SureBlue™ TMB Microwell Peroxidase Substrate, KPL Inc.) was added and allowed to react for 10 min. The reaction was stopped with TMB-Stop Solution (KPL Inc.), and the optical density (OD) was determined at 450 nm – 620 nm using an automated spectrophotometer (PHOmo, Autobio Labtec Instruments). For the serum samples, arbitrary ELISA units (AEU/mL) were calculated by generating a 4-parameter (4PL) logistical fit of the OD values of the standards. AEU/mL were converted into binding antibody units (BAU/mL) relative to the international standard of the WHO (S1) by a factor of 0.07065. The results of the BAL samples were reported as BAU values.

Anti-NCP-IgG titers were determined with the Anti-SARS-CoV-2-NCP-ELISA (IgG) following the manufacturer’s instructions (#EI 2606-9601-2 G, Euroimmun). The NCP ratio corresponds to the absorbance of the sample divided through the absorbance of the reference calibrator. Ratio < 0.8: seronegative; Ratio ≥ 0.8 to < 1.1: borderline; Ratio ≥ 1.1: seropositive. Optical densities were acquired with SpectraMax iD3 (Molecular Devices) and the software SoftMax Pro 7.1.2.

For assessing MVA-specific IgG and IgA, ELISA plates (Nunc MaxiSorp Plates, Thermo Fisher Scientific) were coated with 2x10^6^ plaque-forming units (PFU)/mL MVA LMU F6 (27) overnight at 4°C. After blocking with PBS + 3% nonfat dried milk powder, diluted serum and BAL samples were incubated for 1 h at 37°C. Polyclonal rabbit anti-human IgG HRP (#P0214, Agilent Technologies) or anti-human IgA HRP (#P0216, Agilent Technologies) was added to the plates and incubated for 1 h at 37°C. For detection, TMB substrate solution (#T0440-100ML, Merck) was added and the reaction terminated with stop reagent (#S5814-100ML, Merck). The results were reported as OD450 values for serum samples diluted 1:128 (for assessing MVA-specific IgG) and for BAL samples diluted 1:2 (for assessing MVA-specific IgA).

### Virus neutralization test

The serum neutralization capacity against SARS-CoV-2 was assessed by VNT_100_ (virus neutralization test) as described previously (28). Briefly, serum samples were heat-inactivated for 30 min at 56 °C and diluted in a two-fold dilution series (1:4–1:512) in 96-well cell culture plates, followed by addition of 100 PFU of SARS-CoV-2 (German isolate BavPat1/2020; European Virus Archive Global #026V-03883 (Genbank: MZ558051.1)). BAL samples were diluted in a two-fold dilution series starting from 1:2. After 1 h of incubation at 37°C, 2 × 10^4^ Vero C1008 cells (ATCC, Cat. no. CRL-1586, RRID: CVCL_0574) were added. Cytopathic effects were evaluated at day 4 post infection. Neutralization was defined as the absence of cytopathic effects, and the reciprocal neutralization titer was calculated from the highest serum dilution without cytopathic effects as a geometric mean based on three replicates. The lower limit of detection is a reciprocal titer of 8 (serum) and 4 (BAL), respectively.

### Interferon-gamma release assay (IGRA)

IFN-γ secretion by S1-specific T cells was analyzed in lithium-heparin whole blood using a IFN-γ release T cell assay (#ET 2606-3003, Euroimmun). After 20-24 h of stimulation, IFN-γ was measured in the plasma using an IFN-γ ELISA (#EQ 6841-9601, Euroimmun) according to manufacturer’s instructions. Optical densities were acquired with SpectraMax iD3 (Molecular Devices) and the software SoftMax Pro 7.1.2.

### SARS-CoV-2 spike peptide pools

We ordered 15 amino acid (aa) long and 10 aa overlapping peptide pools spanning the whole length of Wuhan SARS-CoV-2 spike protein (total 253 peptides) from GenScript. All lyophilized peptides were synthesized at >95% purity and reconstituted at a stock concentration of 50 mg/mL in DMSO (Sigma-Aldrich), except for 9 peptides (number 24, 190, 191, 225, 226, 234, 244, 245, and 246) that were dissolved at 25 mg/mL due to solubility issues. All peptides in DMSO stocks were stored at −80 °C until use.

### T cell re-stimulation assay

PBMCs, isolated using a Ficoll gradient, or BAL cells, isolated by centrifugation of BAL fluid, were re-suspended at a concentration of 2 × 10^7^ cells/mL in complete RPMI medium [RPMI 1640 (Gibco) supplemented with 10% FBS (GE Healthcare Life Sciences), 1 mM sodium pyruvate, 50 µM β-mercaptoethanol, 1% streptomycin/penicillin (all Gibco)]. For stimulation, cells were diluted with the spike peptide pool. The peptide pool was prepared in complete RPMI with or without brefeldin A (Sigma-Aldrich) at a final concentration of 10 µg/mL. In the final mixture, each peptide had a concentration of 2 µg (∼1.2 nmol)/mL, except for above mentioned peptides, which were used at a final concentration of 1 µg/mL due to solubility issues. As a negative control, we stimulated the cells with DMSO, in a volume corresponding to the DMSO amount in peptide pools (equaling to 5 % DMSO in final medium volume). In each experiment, we used cells stimulated with Phorbol-12-myristate-13-acetate (PMA; Calbiochem) and ionomycin (Invitrogen) at final concentration of 50 ng/mL and 1500 ng/mL, respectively, as an internal positive control. Cells were incubated for 16 h at 37 °C, 5% CO_2_. The supernatant of cells stimulated in the absence of brefeldin A was harvested and stored at -80°C for later bead-based cytokine analysis. The cells stimulated in the presence of brefeldin A were washed and resuspended in MACS buffer (PBS, 3% FBS, 2 mM EDTA). Non-specific antibody binding was blocked by incubating samples with 10% mouse serum at 4°C for 15 min. Next, without washing, an antibody mix of anti-CD3-APC-Fire810 (SK7; # 344858; Biolegend; 1:50), anti-CD4-BUV563 (RPA-T4; #741353; BD Biosciences; 1:200), anti-CD8-SparkBlue550 (SK1; #344760; Biolegend; 1:200), anti-CD45RA-BUV395 (HI100, #740298; BD Biosciences; 1:200), anti-CCR7-BV785 (G043H7; #353230; Biolegend; 1:50) and Zombie Yellow™ Fixable Viability Kit (#423104; BioLegend; 1:400) was added. After staining for 20 min at RT, cells were washed before they were fixed and permeabilized (#554714; BD Biosciences) according to the manufacturers’ protocol. Intracellular cytokines were stained using anti-IFN-γ-PE-Cy7 (B27; #506518; Biolegend; 1:100), anti-TNF-α-AF700 (Mab11; #502928; Biolegend; 1:50) for 45 min on RT. Cells were acquired on Cytek Aurora spectral flow cytometer (Cytek) equipped with five lasers operating on 355 nm, 405 nm, 488 nm, 561 nm, and 640 nm. Flow cytometry data were acquired using SpectroFlo v2.2.0 (Cytek) and analyzed by FCS Express V7 (Denovo).

### Bead-based multiplex IgA immunoassay

SARS-CoV-2 S1-, RBD-, S2- and NCP-specific antibodies were detected using the SARS-CoV-2 Antigen Panel 1 IgA assay (Millipore, HC19SERA1-85K-04) following the manufacturer’s instructions. Plasma samples were diluted 1:250, while BAL, bronchosorption and nasosorption samples were diluted 1:5. The readout is given as median fluorescence intensity (MFI) of > 50 beads for each antigen and sample, acquired by the Bio-Plex 200 machine and the Bio-Plex Manager Version 6.0 software (Bio-Rad).

### Multiplex cytokine assay

The supernatants of cells re-stimulated with spike peptides in the absence of brefeldin were examined using a Multiplex cytokine assay (Bio-Plex Pro Human Cytokine Screening Panel 48-Plex, #12007283 Biorad) following the manufacturer’s instructions. Briefly, magnetic capture beads were pipetted into 96-well plates in equal amounts and then washed twice in wash buffer solution. Fifty μL of diluted samples (1:2) and standards were added to the plates and incubated for 30 min at RT. After incubation, the plates were washed three times in wash buffer solution and detection antibodies were pipetted into each well in equal amounts to be incubated for 30 min at RT. After incubation, the plates were once again washed three times and a streptavidin-phycoerythrin solution was added to each well in equal amounts for 10 min incubation at RT. Plates were then washed three times and measured in a Bio-Plex 200 System (Biorad). For analysis, measured concentrations below the lower limit of detection were set to half the minimum value of the standard curve of the respective analyte.

In addition, TNFα was quantified in IGRA supernatants using the LEGENDplex Human Essential Immune Response Panel (Biolegend) according to manufacturer’s instructions. Data were acquired on Cytek Aurora spectral flow cytometer (Cytek) and analyzed with the LEGENDplex Data Analysis Software Suite (Qognit).

### Statistics

The analyses have been conducted with Stata/MP 18.0 (StataCorp) and with GraphPad Prism 8.4 (GraphPad Software). In this trial, it was not intended to test hypotheses in a confirmatory sense. Data were summarized by numbers of non-missing values, means with two-sided 95% confidence interval, medians, standard deviations, coefficients of variance, minimum, maximum and interquartile ranges. Wilcoxon matched-pairs signed-rank tests on equality of pre- and post-boosting measurements (i.e. on the null hypothesis of no change from baseline) were applied. The baseline is defined as the time point closest but prior to the inhalation boost. These tests on change from baseline are of exploratory nature only. P values were therefore not adjusted for multiplicity. The summary statistics are provided in Supplementary Tables S1 through S42.

## Data availability

All the data and methods are presented in the manuscript or in the supplemental materials. All individual values for figures are available in the Supporting Data Values file.

## Author contributions

According to CRediT, Contributor Roles Taxonomy, https://credit.niso.org/

| **Roles** | **Authors/Contributors** |
| --- | --- |
| Conceptualization | JMH, RF |
| Data curation | SIH, LR, AB |
| Formal analysis | SIH, RGJ, JBM, IO, LR, VK, AB, GK, LK, CF |
| Funding acquisition | RF, VK, SB |
| Investigation | SIH, RGJ, JBM, IO, VK, GK, MF, IR, JR, AS, LK |
| Methodology | SIH, RGJ, JBM, IO, LR, VK, AB, GK, AT, AZ, CF, SB, CS, JMH, RF |
| Project administration | SIH, SS, CS, JMH, RF |
| Resources | AT, AZ, CF, SB, GS, AV, CS, JMH, RF |
| Software |  |
| Supervision | JMH, RF |
| Validation | VK, MB |
| Visualization | SIH, RF |
| Writing – original draft | SIH |
| Writing – review & editing | All authors |

## Acknowledgements

The authors thank all volunteers for study participation, IDT Biologika (Dessau) for investigational medicinal product supply and the team members of MHH ZKS and Fraunhofer ITEM at the Clinical Research Center (CRC) Hannover for their excellent work.

