## Supplemental Material for "Inhaled booster vaccination with an MVA-based SARS-CoV-2 vaccine candidate induces T cell responses in lung"

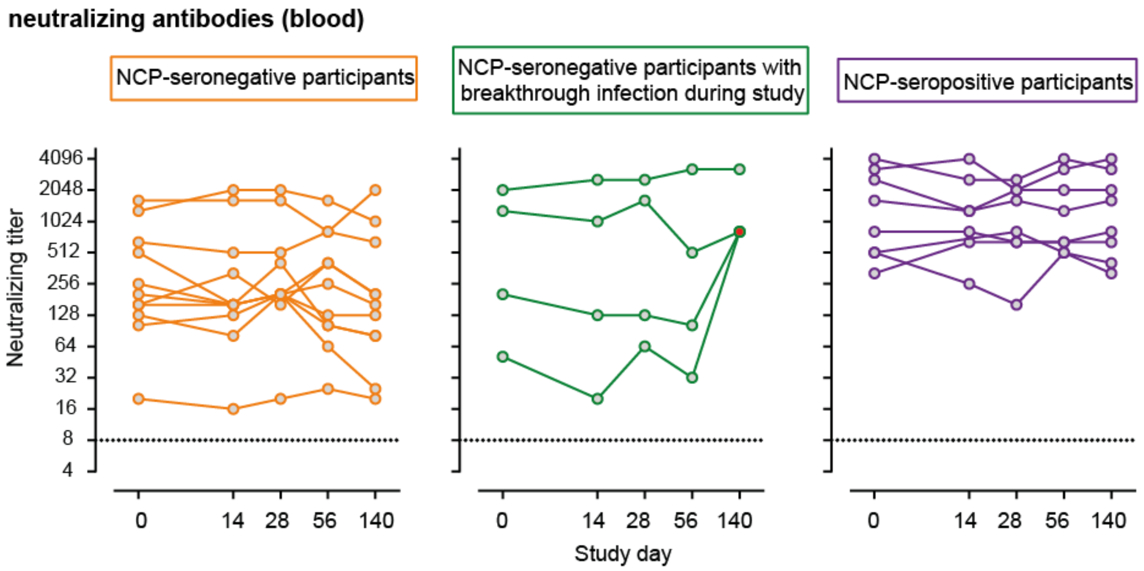


**Supplementary Figure S1**: Analysis of SARS-CoV-2 neutralizing antibodies in blood after MVA-SARS-2-ST inhalation.

Neutralizing activity before and after inhalation in the serum of participants without SARS-CoV-2 breakthrough infection (orange circles), participants with breakthrough infection during the study (green circles), and participants with previous breakthrough infection before the start of the study (purple circles). Red-filled circles represent samples after reported breakthrough infection. The dotted line represents the lower limit of detection of the SARS-CoV-2 $VNT_{100} = 8$.

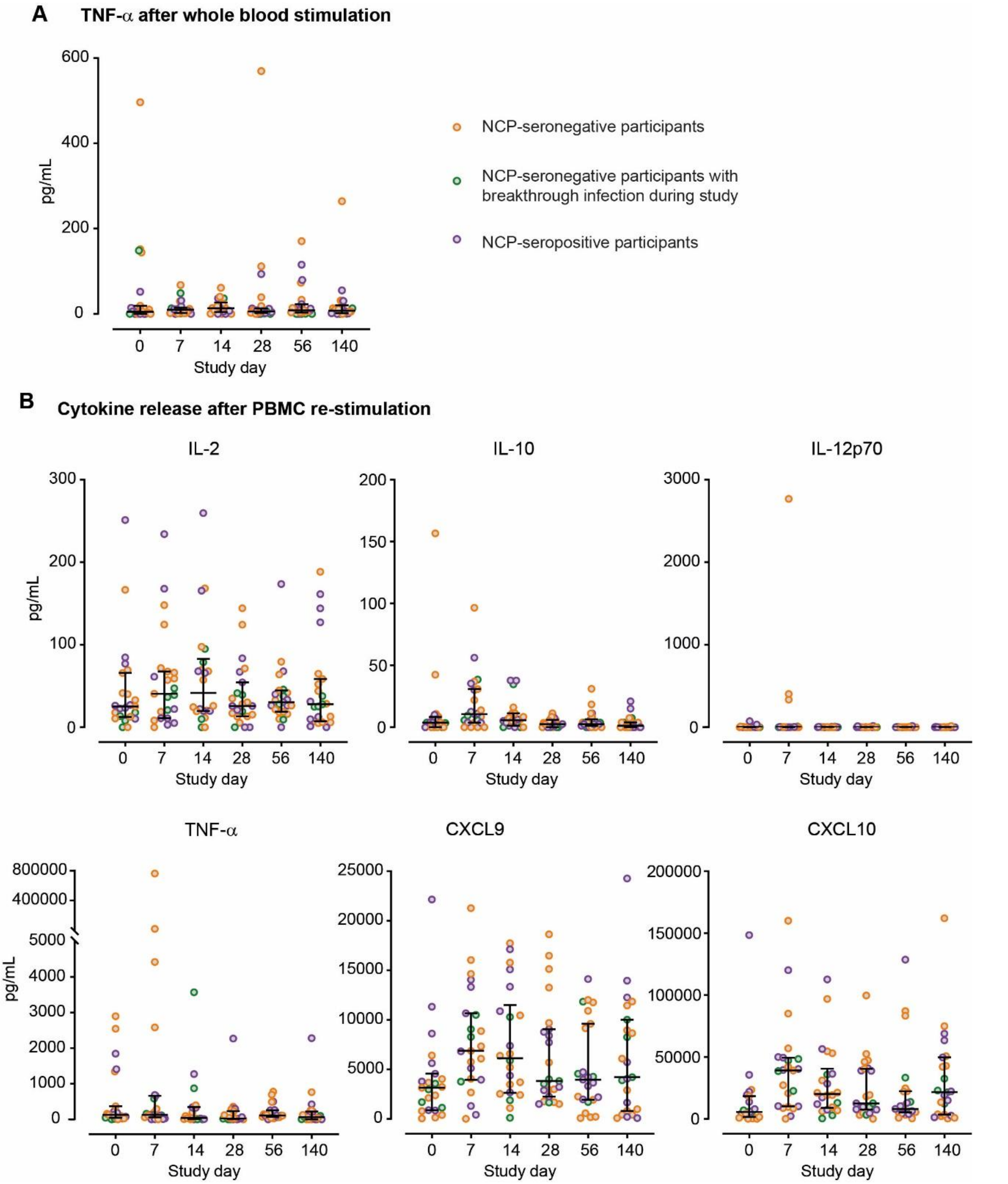


**Supplementary Figure S2**: Cytokine release after whole blood and PBMC re-stimulation with Spike peptides.

(A) TNF-α concentration in full blood supernatants after stimulation with SARS-CoV-2 S1 domain for 20–24 h measured by cytometric bead assay. (B) Cytokine release after re-stimulation of peripheral blood mononuclear cells (PBMC) with spike peptides. (A-B) Lines represent group median with interquartile range; dots represent individual participants: participants without SARS-CoV-2 breakthrough infection (orange circles), participants with breakthrough infection during the study (green circles), and participants with previous breakthrough infection before the start of the study (purple circles). Summary statistics are provided in Supplementary Tables S30 through S36.

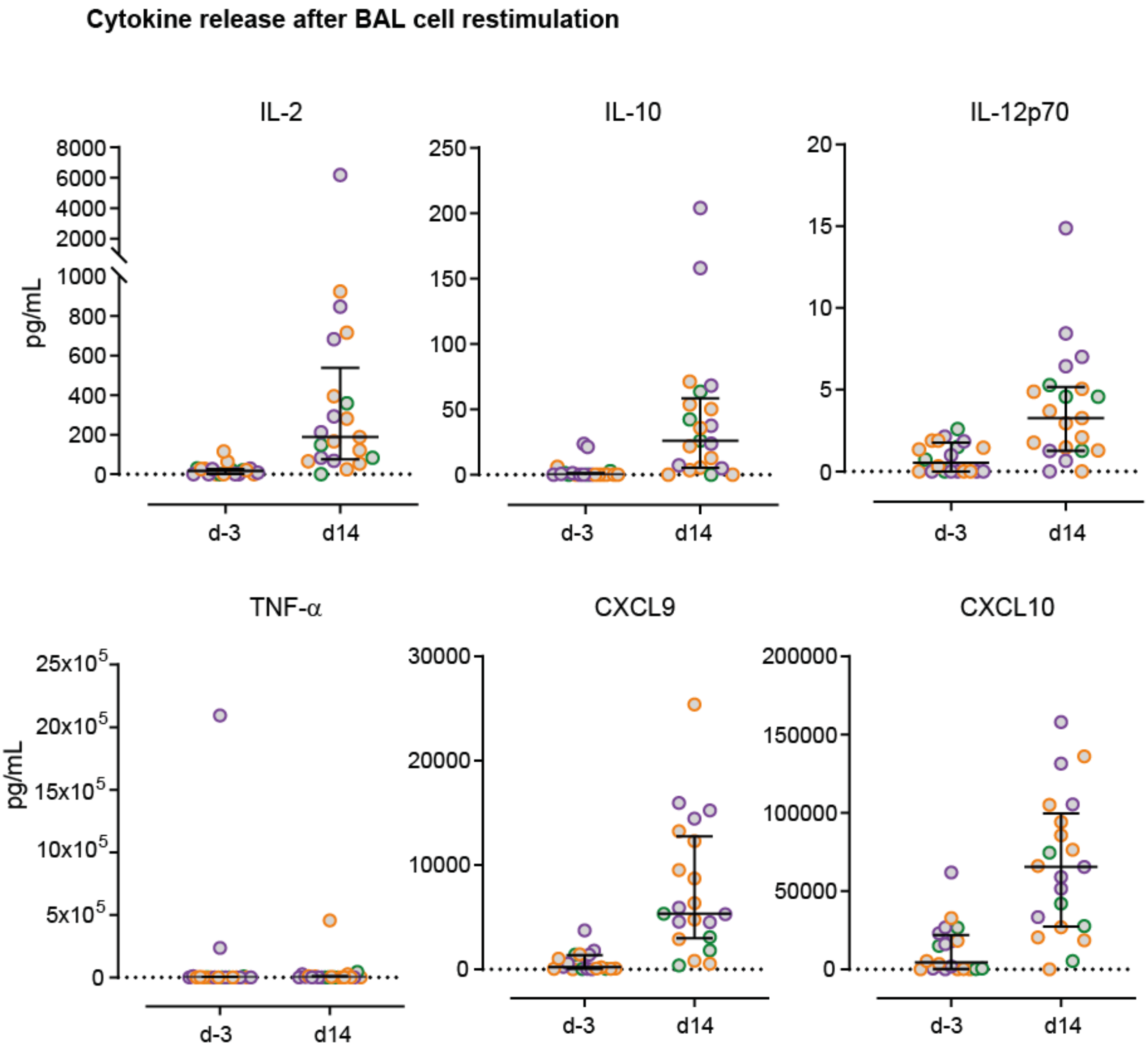


**Supplementary Figure S3**: Cytokine release after re-stimulation of BAL cells with Spike peptides. Cytokine release after re-stimulation of BAL cells with spike peptides. Lines represent group median with interquartile range; dots represent individual participants: participants without SARS-CoV-2 breakthrough infection (orange circles), participants with breakthrough infection during the study (green circles), and participants with previous breakthrough infection before the start of the study (purple circles). Summary statistics are provided in Supplementary Tables S37 through S42.

**Descriptive statistics for Figure 1B:**

**Supplementary Table 1:** Levels of S1-binding IgG antibodies in blood (participants without SARS-CoV-2 breakthrough infection) – absolute values

| S1-binding IgG in blood (BAU/mL) (participants without breakthrough infection) | | | | | | | | |
|---|---|---|---|---|---|---|---|---|
| | n | Mean [95% CI] | SD | CV | [Min, Max] | Median | IQR | p |
| at baseline | 11 | 821.6 [286.7, 1357] | 796.3 | 0.969 | [133, 2670] | 621.0 | 533.0 | |
| at day 7 | 11 | 794.2 [295.9, 1292] | 741.7 | 0.934 | [150, 2222] | 574.0 | 771.0 | 0.700 |
| at day 14 | 11 | 754.2 [284.5, 1224] | 699.2 | 0.927 | [160, 2316] | 553.0 | 679.0 | 0.365 |
| at day 28 | 11 | 733.2 [301.0, 1165] | 643.3 | 0.877 | [205, 2297] | 511.0 | 509.0 | 0.084 |
| at day 56 | 11 | 737.7 [296.5, 1179] | 656.7 | 0.890 | [184, 2158] | 508.0 | 438.0 | 0.083 |
| at day 140 | 11 | 623.6 [242.9, 1004] | 566.7 | 0.909 | [160, 2008] | 402.0 | 399.0 | 0.019 |

n: Number of non-missing values; CI: Confidence interval; SD: Standard Deviation; CV: Coefficient of Variation; Min: Minimum; Max: Maximum; IQR: Interquartile Range. Wilcoxon matched-pairs signed-rank tests on equality of pre- and post-boosting measurements.

**Supplementary Table 2:** Levels of S1-binding IgG antibodies in blood (participants with SARS-CoV-2 breakthrough infection during the study) - absolute values

| S1-binding IgG in blood (BAU/mL) (participants with breakthrough infection during study) | | | | | | | | |
|---|---|---|---|---|---|---|---|---|
| | n | Mean [95% CI] | SD | CV | [Min, Max] | Median | IQR | p |
| at baseline | 4 | 1389.3 [-902.7, 3681] | 1440.4 | 1.037 | [194, 3265] | 1049.0 | 2664.8 | |
| at day 7 | 4 | 1485.5 [-676.1, 3647] | 1358.5 | 0.914 | [192, 2904] | 1423.0 | 2514.0 | 0.875 |
| at day 14 | 4 | 1197.8 [-633.9, 3029] | 1151.1 | 0.961 | [118, 2380] | 1146.5 | 2115.8 | 0.625 |
| at day 28 | 4 | 1311.0 [-727.4, 3349] | 1281.1 | 0.977 | [146, 2761] | 1168.5 | 2383.0 | 0.625 |
| at day 56 | 4 | 1219.0 [-525.6, 2964] | 1096.4 | 0.899 | [155, 2198] | 1261.5 | 1967.0 | 0.999 |
| at day 140 | 4 | 4018.5 [222.3, 7815] | 2385.7 | 0.594 | [2551, 7544] | 2989.5 | 3961.0 | 0.250 |

n: Number of non-missing values; CI: Confidence interval; SD: Standard Deviation; CV: Coefficient of Variation; Min: Minimum; Max: Maximum; IQR: Interquartile Range. Wilcoxon matched-pairs signed-rank tests on equality of pre- and post-boosting measurements.

**Supplementary Table 3:** Levels of S1-binding IgG antibodies in blood (participants with SARS-CoV-2 breakthrough infection before the start of the study) – absolute values

| S1-binding IgG in blood (BAU/mL) (participants with breakthrough infection before study) | | | | | | | | |
|---|---|---|---|---|---|---|---|---|
| | n | Mean [95% CI] | SD | CV | [Min, Max] | Median | IQR | p |
| at baseline | 8 | 2138.4 [1135, 3142] | 1200.4 | 0.561 | [747, 4199] | 1846.0 | 1955.3 | |
| at day 7 | 8 | 2227.6 [1129, 3326] | 1314.3 | 0.590 | [729, 4946] | 1892.5 | 1559.8 | 0.313 |
| at day 14 | 7 | 2602.9 [1238, 3968] | 1475.8 | 0.567 | [1350, 5514] | 2123.0 | 1984.0 | 0.469 |
| at day 28 | 8 | 2675.1 [1328, 4022] | 1611.2 | 0.602 | [846, 5269] | 2051.5 | 2877.8 | 0.078 |
| at day 56 | 8 | 2412.4 [1374, 3451] | 1242.1 | 0.515 | [774, 4854] | 2212.5 | 1497.5 | 0.109 |
| at day 140 | 8 | 2006.4 [1198, 2814] | 966.6 | 0.482 | [701, 3422] | 1946.0 | 1809.0 | 0.461 |

n: Number of non-missing values; CI: Confidence interval; SD: Standard Deviation; CV: Coefficient of Variation; Min: Minimum; Max: Maximum; IQR: Interquartile Range. Wilcoxon matched-pairs signed-rank tests on equality of pre- and post-boosting measurements.

**Descriptive statistics for Figure 1C:**

**Supplementary Table 4:** Levels of S1-binding IgA antibodies in blood (participants without SARS-CoV-2 breakthrough infection) – absolute values

| **S1-binding IgA in blood (MFI) (participants without breakthrough infection)** | | | | | | | | |
|---|---|---|---|---|---|---|---|---|
| | n | Mean [95% CI] | SD | CV | [Min, Max] | Median | IQR | p |
| at baseline | 11 | 296.4 [96.2, 496.7] | 298.0 | 1.005 | [18, 982] | 228.0 | 475.0 | |
| at day 7 | 11 | 289.4 [100.5, 478.3] | 281.2 | 0.971 | [18, 927] | 299.0 | 457.0 | 0.232 |
| at day 14 | 11 | 296.1 [121.0, 471.2] | 260.6 | 0.880 | [16, 816] | 274.0 | 429.0 | 0.700 |
| at day 28 | 11 | 302.8 [113.3, 492.3] | 282.1 | 0.932 | [17, 924] | 274.0 | 438.0 | 0.999 |
| at day 56 | 11 | 301.3 [111.1, 491.4] | 283.0 | 0.939 | [19, 908] | 256.0 | 450.0 | 0.700 |
| at day 140 | 11 | 253.4 [88.1, 418.7] | 246.0 | 0.971 | [14, 785] | 202.0 | 398.5 | 0.148 |

n: Number of non-missing values; CI: Confidence interval; SD: Standard Deviation; CV: Coefficient of Variation; Min: Minimum; Max: Maximum; IQR: Interquartile Range. Wilcoxon matched-pairs signed-rank tests on equality of pre- and post-boosting measurements.

**Supplementary Table 5:** Levels of S1-binding IgA antibodies in blood (participants with SARS-CoV-2 breakthrough infection during the study) – absolute values

| **S1-binding IgA in blood (MFI) (participants with breakthrough infection during study)** | | | | | | | | |
|---|---|---|---|---|---|---|---|---|
| | n | Mean [95% CI] | SD | CV | [Min, Max] | Median | IQR | p |
| at baseline | 4 | 220.1 [-194.1, 634.4] | 260.3 | 1.183 | [22, 579] | 139.8 | 470.6 | |
| at day 7 | 4 | 231.1 [-209.5, 671.7] | 276.9 | 1.198 | [22, 615] | 143.8 | 499.1 | 0.250 |
| at day 14 | 4 | 227.6 [-208.1, 663.4] | 273.8 | 1.203 | [24, 610] | 138.3 | 491.1 | 0.625 |
| at day 28 | 4 | 222.5 [-209.1, 654.1] | 271.2 | 1.219 | [21, 602] | 133.5 | 486.0 | 0.750 |
| at day 56 | 4 | 230.1 [-217.5, 677.8] | 281.3 | 1.222 | [22, 625] | 137.0 | 503.4 | 0.999 |
| at day 140 | 4 | 591.8 [245.9, 937.6] | 217.3 | 0.367 | [292, 804] | 635.5 | 403.3 | 0.125 |

n: Number of non-missing values; CI: Confidence interval; SD: Standard Deviation; CV: Coefficient of Variation; Min: Minimum; Max: Maximum; IQR: Interquartile Range. Wilcoxon matched-pairs signed-rank tests on equality of pre- and post-boosting measurements.

**Supplementary Table 6:** Levels of S1-binding IgA antibodies in blood (participants with SARS-CoV-2 breakthrough infection before the start of the study) – absolute values

| **S1-binding IgA in blood (MFI) (participants with breakthrough infection before study)** | | | | | | | | |
|---|---|---|---|---|---|---|---|---|
| | n | Mean [95% CI] | SD | CV | [Min, Max] | Median | IQR | p |
| at baseline | 8 | 1631.3 [424.2, 2838] | 1443.8 | 0.885 | [164, 4851] | 1216.5 | 1258.3 | |
| at day 7 | 8 | 1694.6 [485.4, 2904] | 1446.4 | 0.854 | [237, 4864] | 1250.3 | 1428.3 | 0.016 |
| at day 14 | 7 | 1945.1 [571.6, 3319] | 1485.2 | 0.764 | [850, 5038] | 1375.0 | 1626.0 | 0.109 |
| at day 28 | 8 | 1717.8 [506.0, 2930] | 1449.4 | 0.844 | [217, 4895] | 1301.3 | 1425.1 | 0.016 |
| at day 56 | 8 | 1707.8 [420.0, 2995] | 1540.3 | 0.902 | [189, 5095] | 1102.5 | 1496.5 | 0.195 |
| at day 140 | 8 | 1628.6 [277.2, 2980] | 1616.5 | 0.993 | [124, 5237] | 1024.0 | 1513.0 | 0.945 |

n: Number of non-missing values; CI: Confidence interval; SD: Standard Deviation; CV: Coefficient of Variation; Min: Minimum; Max: Maximum; IQR: Interquartile Range. Wilcoxon matched-pairs signed-rank tests on equality of pre- and post-boosting measurements.

### Descriptive statistics for Figure 2A:

**Supplementary Table 7**: IFN-gamma (re-stimulation of whole blood) - Absolute values

| IFN-gamma (mIU/mL) | | | | | | | | |
|---|---|---|---|---|---|---|---|---|
| | n | Mean [95% CI] | SD | CV | [Min, Max] | Median | IQR | p |
| at baseline | 23 | 1739.249 [756.143, 2722.355] | 2273.432 | 1.307 | [111.30, 10983.90] | 948.200 | 1492.000 | |
| at day 7 | 23 | 9569.856 [4850.872, 14288.840] | 10912.650 | 1.140 | [970.20, 50344.54] | 5274.200 | 10261.400 | 0.000 |
| at day 14 | 22 | 6396.958 [2637.058, 10156.858] | 8480.177 | 1.326 | [437.50, 39051.40] | 3382.820 | 6336.000 | 0.000 |
| at day 28 | 23 | 3064.014 [1899.939, 4228.090] | 2691.925 | 0.879 | [412.40, 10805.53] | 2252.400 | 2784.000 | 0.001 |
| at day 56 | 23 | 2610.130 [231.000, 19953.25] | 3958.641 | 1.517 | [-270, 322] | 1834.500 | 2017.900 | 0.020 |
| at day 140 | 23 | 2655.639 [1487.987, 3823.292] | 2700.196 | 1.017 | [276.00, 12535.95] | 2345.200 | 1992.600 | 0.016 |

n: Number of non-missing values; CI: Confidence interval; SD: Standard Deviation; CV: Coefficient of Variation; Min: Minimum; Max: Maximum; IQR: Interquartile Range. Wilcoxon matched-pairs signed-rank tests on equality of pre- and post-boosting measurements.

### Descriptive statistics for Figure 2B:

**Supplementary Table 8**: percentage of cytokine-secreting $CD4^+$ T cells after PBMC re-stimulation

| Cytokine-secreting CD4+ T cells, blood (%) | | | | | | | | |
|---|---|---|---|---|---|---|---|---|
| | n | Mean [95% CI] | SD | CV | [Min, Max] | Median | IQR | p |
| at baseline | 23 | 0.062 [0.013, 0.110] | 0.113 | 1.822 | [0.006, 0.526] | 0.029 | 0.038 | |
| at day 7 | 23 | 0.049 [-0.005, 0.103] | 0.124 | 2.531 | [-0.474, 0.207] | 0.054 | 0.077 | 0.105 |
| at day 14 | 22 | 0.067 [0.040, 0.093] | 0.059 | 0.890 | [0.008, 0.207] | 0.041 | 0.049 | 0.106 |
| at day 28 | 23 | 0.075 [0.0195, 0.131] | 0.129 | 1.715 | [-0.018, 0.621] | 0.030 | 0.057 | 0.286 |
| at day 56 | 23 | 0.037 [0.024, 0.051] | 0.032 | 0.853 | [0.000, 0.124] | 0.026 | 0.042 | 0.710 |
| at day 140 | 23 | 0.044 [0.029, 0.060] | 0.036 | 0.822 | [-0.041, 0.120] | 0.037 | 0.046 | 0.273 |

n: Number of non-missing values; CI: Confidence interval; SD: Standard Deviation; CV: Coefficient of Variation; Min: Minimum; Max: Maximum; IQR: Interquartile Range. Wilcoxon matched-pairs signed-rank tests on equality of pre- and post-boosting measurements.

**Supplementary Table 9**: percentage of cytokine-secreting CD8$^+$ T cells after PBMC re-stimulation

| **Cytokine-secreting CD8+ T cells, blood (%)** | | | | | | | | |
|---|---|---|---|---|---|---|---|---|
| | n | Mean [95% CI] | SD | CV | [Min, Max] | Median | IQR | p |
| at baseline | 23 | 0.065 [0.019, 0112] | 0.108 | 1.650 | [-0.030, 0.375] | 0.021 | 0.109 | |
| at day 7 | 23 | 0.070 [-0.014, 0.155] | 0.196 | 2.787 | [-0.218, 0.903] | 0.022 | 0.074 | 0.870 |
| at day 14 | 22 | 0.095 [-0.003, 0.192] | 0.220 | 2.327 | [-0.001, 1.048] | 0.021 | 0.107 | 0.503 |
| at day 28 | 23 | 0.107 [-0.001, 0.215] | 0.249 | 2.322 | [-0.030, 0.946] | 0.028 | 0.066 | 0.777 |
| at day 56 | 23 | 0.039 [0.008, 0.069] | 0.070 | 1.813 | [-0.008, 0.348] | 0.026 | 0.036 | 0.496 |
| at day 140 | 23 | 0.084 [-0.005, 0.174] | 0.206 | 2.443 | [-0.120, 0.972] | 0.033 | 0.054 | 0.777 |

n: Number of non-missing values; CI: Confidence interval; SD: Standard Deviation; CV: Coefficient of Variation; Min: Minimum; Max: Maximum; IQR: Interquartile Range. Wilcoxon matched-pairs signed-rank tests on equality of pre- and post-boosting measurements.

**<u>Descriptive statistics for Figure 2C:</u>**

**Supplementary Table 10**: IFN-gamma release after PBMC re-stimulation - Absolute values

| **IFN-gamma (pg/mL)** | | | | | | | | |
|---|---|---|---|---|---|---|---|---|
| | n | Mean [95% CI] | SD | CV | [Min, Max] | Median | IQR | p |
| at baseline | 23 | 65.067 [-8.172, 138.307] | 169.366 | 2.603 | [0.00, 819.36] | 20.260 | 28.620 | |
| at day 7 | 23 | 584.880 [-58.140, 1227.900 | 1486.983 | 2.542 | [0.00, 6832.70] | 78.740 | 340.520 | 0.002 |
| at day 14 | 22 | 185.793 [-36.458, 408.044] | 501.271 | 2.698 | [0.00, 2375.12] | 24.025 | 129.280 | 0.187 |
| at day 28 | 23 | 113.658 [10.416, 216.900] | 238.747 | 2.101 | [0.00, 1073.64] | 23.780 | 96.850 | 0.273 |
| at day 56 | 23 | 115.330 [231.000, 19953.25] | 337.424 | 2.926 | [0.00, 1614.020] | 22.090 | 31.870 | 0.560 |
| at day 140 | 23 | 49.135 [19.003, 79.267] | 69.680 | 1.418 | [0.00, 218.79] | 17.600 | 46.950 | 0.893 |

n: Number of non-missing values; CI: Confidence interval; SD: Standard Deviation; CV: Coefficient of Variation; Min: Minimum; Max: Maximum; IQR: Interquartile Range. Wilcoxon matched-pairs signed-rank tests on equality of pre- and post-boosting measurements.

**Supplementary Table 11**: IL-4 release after PBMC re-stimulation - Absolute values

| IL-4 (pg/mL) | | | | | | | | |
|---|---|---|---|---|---|---|---|---|
| | n | Mean [95% CI] | SD | CV | [Min, Max] | Median | IQR | p |
| at baseline | 23 | 4.814 [2.656, 6.973] | 4.991 | 1.037 | [0.00, 21.92] | 4.37 | 6.33 | |
| at day 7 | 23 | 3.088 [1.545, 4.631] | 3.568 | 1.155 | [0.00, 10.95] | 1.7 | 5.19 | 0.322 |
| at day 14 | 22 | 3.280 [1.403, 5.157] | 4.233 | 1.29 | [0.00, 16.69] | 1.69 | 7.06 | 0.345 |
| at day 28 | 23 | 1.874 [0.960, 2.788] | 2.113 | 1.128 | [0.00, 8.37] | 1.51 | 3.12 | 0.016 |
| at day 56 | 23 | 4.186 [2.331, 6.041] | 4.290 | 1.025 | [0.00, 15.79] | 2.74 | 6.02 | 0.33 |
| at day 140 | 23 | 4.356 [2.137, 6.574] | 5.129 | 1.178 | [0.00, 20.26] | 3.57 | 6.02 | 0.964 |

n: Number of non-missing values; CI: Confidence interval; SD: Standard Deviation; CV: Coefficient of Variation; Min: Minimum; Max: Maximum; IQR: Interquartile Range. Wilcoxon matched-pairs signed-rank tests on equality of pre- and post-boosting measurements.

**Supplementary Table 12**: IL-17 release after PBMC re-stimulation - Absolute values

| IL-17 (pg/mL) | | | | | | | | |
|---|---|---|---|---|---|---|---|---|
| | n | Mean [95% CI] | SD | CV | [Min, Max] | Median | IQR | p |
| at baseline | 23 | 20.847 [9.253, 32.440] | 26.809 | 1.286 | [0.00, 120.40] | 15.61 | 30.11 | |
| at day 7 | 23 | 14.650 [5.180, 24.120] | 21.899 | 1.495 | [0.00, 85.02] | 7.62 | 22.04 | 0.376 |
| at day 14 | 22 | 13.735 [3.821, 23.650] | 22.361 | 1.628 | [0.00, 85.43] | 1.795 | 22.28 | 0.234 |
| at day 28 | 23 | 7.433 [2.806, 12.061] | 10.701 | 1.44 | [0.00, 45.93] | 3.25 | 10.94 | 0.010 |
| at day 56 | 23 | 15.100 [6.706, 23.490] | 5.132 | 1.183 | [0.00, 20.26] | 3.57 | 6.02 | 0.439 |
| at day 140 | 23 | 14.050 [3.884, 24.216] | 23.508 | 1.673 | [0.00, 114.40] | 8.19 | 18.35 | 0.217 |

n: Number of non-missing values; CI: Confidence interval; SD: Standard Deviation; CV: Coefficient of Variation; Min: Minimum; Max: Maximum; IQR: Interquartile Range. Wilcoxon matched-pairs signed-rank tests on equality of pre- and post-boosting measurements.

**Descriptive statistics for Figure 3A:**

**Supplementary Table 13:** Levels of S1-binding IgG antibodies in BAL fluid (participants without SARS-CoV-2 breakthrough infection) – absolute values

| **S1-binding IgG (BAU) in BAL fluid (participants without breakthrough infection)** | | | | | | | | |
|---|---|---|---|---|---|---|---|---|
| | n | Mean [95% CI] | SD | CV | [Min, Max] | Median | IQR | p |
| at baseline | 11 | 18.9 [8.3, 29.2] | 15.6 | 0.826 | [4.0, 53.0] | 14.0 | 27.0 | |
| at day 14 | 11 | 27.5 [6.2, 48.3] | 31.4 | 1.145 | [4.0, 114.0] | 18.0 | 16.0 | 0.168 |

n: Number of non-missing values; CI: Confidence interval; SD: Standard Deviation; CV: Coefficient of Variation; Min: Minimum; Max: Maximum; IQR: Interquartile Range. Wilcoxon matched-pairs signed-rank tests on equality of pre- and post-boosting measurements.

**Supplementary Table 14:** Levels of S1-binding IgG antibodies in BAL fluid (participants with SARS-CoV-2 breakthrough infection during the study) - absolute values

| **S1-binding IgG (BAU) in BAL fluid (participants with breakthrough infection during study)** | | | | | | | | |
|---|---|---|---|---|---|---|---|---|
| | n | Mean [95% CI] | SD | CV | [Min, Max] | Median | IQR | p |
| at baseline | 4 | 50.0 [-29.3, 128.8] | 49.6 | 0.992 | [4.0, 110.0] | 43.0 | 93.5 | |
| at day 14 | 4 | 110.5 [-90.8, 311.1] | 126.4 | 1.144 | [4.0, 271.0] | 83.5 | 234.5 | 0.500 |

n: Number of non-missing values; CI: Confidence interval; SD: Standard Deviation; CV: Coefficient of Variation; Min: Minimum; Max: Maximum; IQR: Interquartile Range. Wilcoxon matched-pairs signed-rank tests on equality of pre- and post-boosting measurements.

**Supplementary Table 15:** Levels of S1-binding IgG antibodies in BAL fluid (participants with SARS-CoV-2 breakthrough infection before the start of the study) – absolute values

| **S1-binding IgG (BAU) in BAL fluid (participants with breakthrough infection before study)** | | | | | | | | |
|---|---|---|---|---|---|---|---|---|
| | n | Mean [95% CI] | SD | CV | [Min, Max] | Median | IQR | p |
| at baseline | 8 | 181.8 [21.8, 341.2] | 191.1 | 1.052 | [25.0, 580.0] | 115.0 | 262.3 | |
| at day 14 | 7 | 174.9 [41.4, 307.9] | 144.0 | 0.824 | [52.0, 398.0] | 121.0 | 305.0 | 0.688 |

n: Number of non-missing values; CI: Confidence interval; SD: Standard Deviation; CV: Coefficient of Variation; Min: Minimum; Max: Maximum; IQR: Interquartile Range. Wilcoxon matched-pairs signed-rank tests on equality of pre- and post-boosting measurements.

**Descriptive statistics for Figure 3B:**

**Supplementary Table 16:** Levels of S1-binding IgA antibodies in BAL fluid (participants without SARS-CoV-2 breakthrough infection) – absolute values

| **S1-binding IgA (MFI) in BAL fluid (participants without breakthrough infection)** | | | | | | | | |
|---|---|---|---|---|---|---|---|---|
| | n | Mean [95% CI] | SD | CV | [Min, Max] | Median | IQR | p |
| at baseline | 11 | 24.1 [7.0, 41.3] | 25.5 | 1.056 | [6.0, 93.0] | 14.0 | 25.0 | |
| at day 14 | 11 | 27.8 [13.7, 42.0] | 21.1 | 0.757 | [7.0, 69.5] | 18.0 | 29.0 | 0.375 |

n: Number of non-missing values; CI: Confidence interval; SD: Standard Deviation; CV: Coefficient of Variation; Min: Minimum; Max: Maximum; IQR: Interquartile Range. Wilcoxon matched-pairs signed-rank tests on equality of pre- and post-boosting measurements.

**Supplementary Table 17:** Levels of S1-binding IgA antibodies in BAL fluid (participants with SARS-CoV-2 breakthrough infection during the study) - absolute values

| **S1-binding IgA (MFI) in BAL fluid (participants with breakthrough infection during study)** | | | | | | | | |
|---|---|---|---|---|---|---|---|---|
| | n | Mean [95% CI] | SD | CV | [Min, Max] | Median | IQR | p |
| at baseline | 4 | 48.8 [-69.7, 167.2] | 74.5 | 1.527 | [6.0, 160.0] | 43.0 | 118.8 | |
| at day 14 | 4 | 57.9 [-45.9, 161.6] | 65.2 | 1.127 | [8.0, 146.0] | 83.5 | 118.4 | 0.625 |

n: Number of non-missing values; CI: Confidence interval; SD: Standard Deviation; CV: Coefficient of Variation; Min: Minimum; Max: Maximum; IQR: Interquartile Range. Wilcoxon matched-pairs signed-rank tests on equality of pre- and post-boosting measurements.

**Supplementary Table 18:** Levels of S1-binding IgA antibodies in BAL fluid (participants with SARS-CoV-2 breakthrough infection before the start of the study) – absolute values

| **S1-binding IgA (MFI) in BAL fluid (participants with breakthrough infection before study)** | | | | | | | | |
|---|---|---|---|---|---|---|---|---|
| | n | Mean [95% CI] | SD | CV | [Min, Max] | Median | IQR | p |
| at baseline | 8 | 252.4 [87.9, 417.0] | 196.8 | 0.780 | [44.0, 535.0] | 231.0 | 404.9 | |
| at day 14 | 7 | 398.5 [176.7, 620.3] | 239.8 | 0.602 | [71.0, 750.0] | 324.5 | 423.0 | 0.063 |

n: Number of non-missing values; CI: Confidence interval; SD: Standard Deviation; CV: Coefficient of Variation; Min: Minimum; Max: Maximum; IQR: Interquartile Range. Wilcoxon matched-pairs signed-rank tests on equality of pre- and post-boosting measurements.

**Descriptive statistics for Figure 3C:**

**Supplementary Table 19:** Levels of MVA-binding IgA antibodies in BAL fluid (participants without SARS-CoV-2 breakthrough infection) – absolute values

| **MVA-binding IgA (OD450) in BAL fluid (participants without breakthrough infection)** | | | | | | | | |
|---|---|---|---|---|---|---|---|---|
| | n | Mean [95% CI] | SD | CV | [Min, Max] | Median | IQR | p |
| at baseline | 11 | 0.031 [0.016, 0.045] | 0.021 | 0.699 | [0.004, 0.075] | 0.026 | 0.031 | |
| at day 14 | 11 | 0.036 [0.013, 0.059] | 0.034 | 0.946 | [0.004, 0.075] | 0.031 | 0.047 | 0.815 |

n: Number of non-missing values; CI: Confidence interval; SD: Standard Deviation; CV: Coefficient of Variation; Min: Minimum; Max: Maximum; IQR: Interquartile Range. Wilcoxon matched-pairs signed-rank tests on equality of pre- and post-boosting measurements.

**Supplementary Table 20:** Levels of MVA-binding IgA antibodies in BAL fluid (participants with SARS-CoV-2 breakthrough infection during the study) - absolute values

| **MVA-binding IgA (OD450) in BAL fluid (participants with breakthrough infection during study)** | | | | | | | | |
|---|---|---|---|---|---|---|---|---|
| | n | Mean [95% CI] | SD | CV | [Min, Max] | Median | IQR | p |
| at baseline | 4 | 0.030 [-0.002, 0.062] | 0.020 | 0.676 | [0.007, 0.056] | 0.029 | 0.039 | |
| at day 14 | 4 | 0.025 [0.000, 0.050] | 0.016 | 0.629 | [0.016, 0.048] | 0.018 | 0.025 | 0.875 |

n: Number of non-missing values; CI: Confidence interval; SD: Standard Deviation; CV: Coefficient of Variation; Min: Minimum; Max: Maximum; IQR: Interquartile Range. Wilcoxon matched-pairs signed-rank tests on equality of pre- and post-boosting measurements.

**Supplementary Table 21:** Levels of MVA-binding IgA antibodies in BAL fluid (participants with SARS-CoV-2 breakthrough infection before the start of the study) – absolute values

| **MVA-binding IgA (OD450) in BAL fluid (participants with breakthrough infection before study)** | | | | | | | | |
|---|---|---|---|---|---|---|---|---|
| | n | Mean [95% CI] | SD | CV | [Min, Max] | Median | IQR | p |
| at baseline | 8 | 0.030 [0.005, 0.054] | 0.029 | 0.976 | [0.008, 0.098] | 0.020 | 0.017 | |
| at day 14 | 7 | 0.055 [0.020, 0.089] | 0.038 | 0.687 | [0.023, 0.136] | 0.046 | 0.026 | 0.313 |

n: Number of non-missing values; CI: Confidence interval; SD: Standard Deviation; CV: Coefficient of Variation; Min: Minimum; Max: Maximum; IQR: Interquartile Range. Wilcoxon matched-pairs signed-rank tests on equality of pre- and post-boosting measurements.

**Descriptive statistics for Figure 3D:**

**Supplementary Table 22:** Levels of S1-binding IgA antibodies in bronchosorption (participants without SARS-CoV-2 breakthrough infection) – absolute values

| **S1-binding IgA (MFI) in bronchosorption (participants without breakthrough infection)** | | | | | | | | |
|---|---|---|---|---|---|---|---|---|
| | n | Mean [95% CI] | SD | CV | [Min, Max] | Median | IQR | p |
| at baseline | 8 | 120.0 [-4.6, 244.6] | 149.0 | 1.242 | [6.0, 373.0] | 61.5 | 263.8 | |
| at day 14 | 9 | 137.7 [-15.1, 290.4] | 198.7 | 1.443 | [6.0, 631.0] | 71.0 | 162.0 | 0.250 |

n: Number of non-missing values; CI: Confidence interval; SD: Standard Deviation; CV: Coefficient of Variation; Min: Minimum; Max: Maximum; IQR: Interquartile Range. Wilcoxon matched-pairs signed-rank tests on equality of pre- and post-boosting measurements.

**Supplementary Table 23:** Levels of S1-binding IgA antibodies in bronchosorption (participants with SARS-CoV-2 breakthrough infection during the study) - absolute values

| **S1-binding IgA (MFI) in bronchosorption (participants with breakthrough infection during study)** | | | | | | | | |
|---|---|---|---|---|---|---|---|---|
| | n | Mean [95% CI] | SD | CV | [Min, Max] | Median | IQR | p |
| at baseline | 3 | 98.5 [-178.1, 375.1] | 111.3 | 1.130 | [8.5, 223.0] | 64.0 | 214.5 | |
| at day 14 | 4 | 92.3 [-156.1, 340.6] | 156.1 | 1.692 | [8.0, 326.0] | 17.5 | 243.3 | 0.999 |

n: Number of non-missing values; CI: Confidence interval; SD: Standard Deviation; CV: Coefficient of Variation; Min: Minimum; Max: Maximum; IQR: Interquartile Range. Wilcoxon matched-pairs signed-rank tests on equality of pre- and post-boosting measurements.

**Supplementary Table 24:** Levels of S1-binding IgA antibodies in bronchosorption (participants with SARS-CoV-2 breakthrough infection before the start of the study) – absolute values

| **S1-binding IgA (MFI) in bronchosorption (participants with breakthrough infection before study)** | | | | | | | | |
|---|---|---|---|---|---|---|---|---|
| | n | Mean [95% CI] | SD | CV | [Min, Max] | Median | IQR | p |
| at baseline | 8 | 1162.0 [47.7, 2277.0] | 1333.0 | 1.147 | [32.0, 3552.0] | 492.0 | 2195.6 | |
| at day 14 | 7 | 2013.0 [-114.5, 4141.3] | 2301.0 | 1.143 | [281.0, 6891.0] | 1832.0 | 1961.0 | 0.813 |

n: Number of non-missing values; CI: Confidence interval; SD: Standard Deviation; CV: Coefficient of Variation; Min: Minimum; Max: Maximum; IQR: Interquartile Range. Wilcoxon matched-pairs signed-rank tests on equality of pre- and post-boosting measurements.

### Descriptive statistics for Figure 4A:

**Supplementary Table 25**: percentage of cytokine-secreting CD4$^+$ T cells after BAL cell re-stimulation

| Cytokine-secreting CD4+ T cells, BAL (%) | | | | | | | | |
|---|---|---|---|---|---|---|---|---|
| | n | Mean [95% CI] | SD | CV | [Min, Max] | Median | IQR | p |
| at baseline | 20 | 0.049 [0.017, 0.081] | 0.069 | 1.410 | [-0.090, 0.210] | 0.050 | 0.074 | |
| at day 14 | 22 | 1.343 [0.773, 1.912] | 1.285 | 0.957 | [0.160, 4.806] | 0.718 | 2.003 | <0.001 |

n: Number of non-missing values; CI: Confidence interval; SD: Standard Deviation; CV: Coefficient of Variation; Min: Minimum; Max: Maximum; IQR: Interquartile Range. Wilcoxon matched-pairs signed-rank tests on equality of pre- and post-boosting measurements.

**Supplementary Table 26**: percentage of cytokine-secreting CD8$^+$ T cells after BAL cell re-stimulation

| Cytokine-secreting CD8+ T cells, BAL (%) | | | | | | | | |
|---|---|---|---|---|---|---|---|---|
| | n | Mean [95% CI] | SD | CV | [Min, Max] | Median | IQR | p |
| at baseline | 20 | 0.061 [-0.042, 0.164] | 0.219 | 3.597 | [-0.057, 0.018] | 0.008 | 0.041 | |
| at day 14 | 22 | 3.456 [1.008, 5.903] | 5.520 | 1.597 | [0.979, 24.524] | 1.354 | 3.845 | <0.001 |

n: Number of non-missing values; CI: Confidence interval; SD: Standard Deviation; CV: Coefficient of Variation; Min: Minimum; Max: Maximum; IQR: Interquartile Range. Wilcoxon matched-pairs signed-rank tests on equality of pre- and post-boosting measurements.

### Descriptive statistics for Figure 4B:

**Supplementary Table 27**: IFN-gamma release after BAL cell re-stimulation - Absolute values

| IFN-gamma (pg/mL) | | | | | | | | |
|---|---|---|---|---|---|---|---|---|
| | n | Mean [95% CI] | SD | CV | [Min, Max] | Median | IQR | p |
| at baseline | 20 | 10.85 [5.04, 16.65] | 12.40 | 1.143 | [0.00, 40.15] | 5.95 | 21.66 | |
| at day 14 | 21 | 2074 [-186, 4333] | 4962.81 | 2.393 | [2.33, 21946.46] | 209.64 | 1567.99 | 0.001 |

n: Number of non-missing values; CI: Confidence interval; SD: Standard Deviation; CV: Coefficient of Variation; Min: Minimum; Max: Maximum; IQR: Interquartile Range. Wilcoxon matched-pairs signed-rank tests on equality of pre- and post-boosting measurements.

**Supplementary Table 28**: IL-4 release after BAL cell re-stimulation - Absolute values

| IL-4 (pg/mL) | | | | | | | | |
|---|---|---|---|---|---|---|---|---|
| | n | Mean [95% CI] | SD | CV | [Min, Max] | Median | IQR | p |
| at baseline | 20 | 0.90 [0.36, 1.44] | 1.16 | 1.280 | [0.00, 4.71] | 0.63 | 1.56 | |
| at day 14 | 21 | 2.63 [1.68, 3.58] | 2.08 | 0.791 | [0.00, 6.73] | 2.33 | 3.91 | 0.002 |

n: Number of non-missing values; CI: Confidence interval; SD: Standard Deviation; CV: Coefficient of Variation; Min: Minimum; Max: Maximum; IQR: Interquartile Range. Wilcoxon matched-pairs signed-rank tests on equality of pre- and post-boosting measurements.

**Supplementary Table 29**: IL-17 release after BAL cell re-stimulation - Absolute values

| IL-17 (pg/mL) | | | | | | | | |
|---|---|---|---|---|---|---|---|---|
| | n | Mean [95% CI] | SD | CV | [Min, Max] | Median | IQR | p |
| at baseline | 20 | 2.90 [0.61, 5.19] | 4.89 | 1.685 | [0.00, 16.46] | 0.00 | 4.08 | |
| at day 14 | 21 | 21.31 [11.73, 30.89] | 21.04 | 0.987 | [0.00, 88.27] | 18.53 | 21.03 | 0.001 |

n: Number of non-missing values; CI: Confidence interval; SD: Standard Deviation; CV: Coefficient of Variation; Min: Minimum; Max: Maximum; IQR: Interquartile Range. Wilcoxon matched-pairs signed-rank tests on equality of pre- and post-boosting measurements.

### Descriptive statistics for Figure S2A:

**Supplementary Table 30**: TNF-alpha (re-stimulation of whole blood) - Absolute values

| TNF-alpha (pg/mL) | | | | | | | | |
|---|---|---|---|---|---|---|---|---|
| | n | Mean [95% CI] | SD | CV | [Min, Max] | Median | IQR | p |
| at baseline | 23 | 46.487 [-1.062, 94.037] | 109.958 | 2.365 | [0.00, 496.25] | 5.18 | 18.96 | |
| at day 7 | 23 | 13.070 [5.767, 20.372] | 16.887 | 1.292 | [0.00, 67.90] | 9.65 | 12.54 | 0.811 |
| at day 14 | 22 | 16.708 [9.513, 23.902] | 16.226 | 0.971 | [0.00, 61.16] | 13.44 | 18.25 | 0.937 |
| at day 28 | 23 | 39.937 [-11.488, 91.362] | 118.921 | 2.978 | [0.00, 569.23] | 5.70 | 10.92 | 0.823 |
| at day 56 | 23 | 25.493 [6.686, 44.300] | 43.493 | 1.706 | [0.00, 170.61] | 8.39 | 19.32 | 0.993 |
| at day 140 | 23 | 22.965 [-0.524, 46.454] | 54.319 | 2.365 | [0.00, 264.16] | 7.31 | 18.48 | 0.743 |

n: Number of non-missing values; CI: Confidence interval; SD: Standard Deviation; CV: Coefficient of Variation; Min: Minimum; Max: Maximum; IQR: Interquartile Range. Wilcoxon matched-pairs signed-rank tests on equality of pre- and post-boosting measurements.

### Descriptive statistics for Figure S2B:

**Supplementary Table 31**: IL-2 release after PBMC re-stimulation - Absolute values

| IL-2 (pg/mL) | | | | | | | | |
|---|---|---|---|---|---|---|---|---|
| | n | Mean [95% CI] | SD | CV | [Min, Max] | Median | IQR | p |
| at baseline | 23 | 45.431 [20.284, 70.578] | 58.152 | 1.280 | [0.00, 251.12] | 25.22 | 53.13 | |
| at day 7 | 23 | 57.474 [31.814, 83.135] | 59.340 | 1.032 | [0.00, 234.10] | 40.67 | 56.63 | 0.111 |
| at day 14 | 22 | 62.139 [33.417, 90.861] | 64.781 | 1.043 | [0.00, 259.51] | 41.68 | 58.98 | 0.156 |
| at day 28 | 23 | 36.965 [20.187, 53.744] | 38.800 | 1.050 | [0.00, 144.72] | 25.75 | 41.51 | 0.731 |
| at day 56 | 23 | 38.808 [23.670, 53.940] | 35.000 | 1.183 | [0.00, 173.59] | 30.26 | 25.80 | 0.482 |
| at day 140 | 23 | 46.875 [23.172, 70.578] | 54.813 | 1.169 | [0.00, 188.39] | 27.99 | 51.32 | 0.893 |

n: Number of non-missing values; CI: Confidence interval; SD: Standard Deviation; CV: Coefficient of Variation; Min: Minimum; Max: Maximum; IQR: Interquartile Range. Wilcoxon matched-pairs signed-rank tests on equality of pre- and post-boosting measurements.

**Supplementary Table 32**: IL-10 release after PBMC re-stimulation - Absolute values

| IL-10 (pg/mL) | | | | | | | | |
|---|---|---|---|---|---|---|---|---|
| | n | Mean [95% CI] | SD | CV | [Min, Max] | Median | IQR | p |
| at baseline | 23 | 12.189 [-1.943, 26.320] | 32.679 | 2.681 | [0.00, 156.61] | 3.75 | 8.49 | |
| at day 7 | 23 | 18.943 [9.038, 28.848] | 22.906 | 1.209 | [0.00, 96.53] | 10.62 | 27.14 | 0.086 |
| at day 14 | 22 | 9.530 [4.282, 14.779] | 11.839 | 1.242 | [0.00, 37.88] | 5.73 | 9.98 | 0.513 |
| at day 28 | 23 | 2.889 [1.513, 4.265] | 3.182 | 1.101 | [0.00, 11.37] | 1.93 | 5.15 | 0.231 |
| at day 56 | 23 | 5.121 [1.975, 8.266] | 7.274 | 1.420 | [0.00, 31.12] | 2.570 | 5.16 | 0.406 |
| at day 140 | 23 | 3.411 [1.134, 5.688] | 5.265 | 1.544 | [0.00, 21.00] | 1.29 | 3.91 | 0.454 |

n: Number of non-missing values; CI: Confidence interval; SD: Standard Deviation; CV: Coefficient of Variation; Min: Minimum; Max: Maximum; IQR: Interquartile Range. Wilcoxon matched-pairs signed-rank tests on equality of pre- and post-boosting measurements.

**Supplementary Table 33**: IL-12p70 release after PBMC re-stimulation - Absolute values

| IL-12p70 (pg/mL) | | | | | | | | |
|---|---|---|---|---|---|---|---|---|
| | n | Mean [95% CI] | SD | CV | [Min, Max] | Median | IQR | p |
| at baseline | 23 | 6.562 [-0.436, 13.559] | 16.182 | 2.466 | [0.00, 73.70] | 1.55 | 3.38 | |
| at day 7 | 23 | 154.147 [-96.379, 404.674] | 579.342 | 3.758 | [0.00, 2766.81] | 1.54 | 5.2 | 0.852 |
| at day 14 | 22 | 2.210 [1.096, 3.324] | 2.513 | 1.137 | [0.00, 8.28] | 1.11 | 3.66 | 0.397 |
| at day 28 | 23 | 2.245 [0.395, 4.095] | 4.279 | 1.906 | [0.00, 16.81] | 0.54 | 2.61 | 0.055 |
| at day 56 | 23 | 2.245 [0.681, 3.810] | 3.618 | 1.611 | [0.00, 17.14] | 1.11 | 2.75 | 0.115 |
| at day 140 | 23 | 1.873 [0.442, 3.303] | 3.308 | 1.767 | [0.00, 15.15] | 1.25 | 2.04 | 0.237 |

n: Number of non-missing values; CI: Confidence interval; SD: Standard Deviation; CV: Coefficient of Variation; Min: Minimum; Max: Maximum; IQR: Interquartile Range. Wilcoxon matched-pairs signed-rank tests on equality of pre- and post-boosting measurements.

**Supplementary Table 34**: TNF-alpha release after PBMC re-stimulation - Absolute values

| TNF-alpha (pg/mL) | | | | | | | | |
|---|---|---|---|---|---|---|---|---|
| | n | Mean [95% CI] | SD | CV | [Min, Max] | Median | IQR | p |
| at baseline | 23 | 522.066 [151.404, 892.729] | 857.157 | 1.642 | [0.00, 2894.14] | 127.81 | 320.26 | |
| at day 7 | 23 | 34548.316 [-3.42e+04, 1.03e+05] | 1.59E+05 | 4.599 | [0.00, 763179.19] | 128.37 | 606.05 | 0.846 |
| at day 14 | 22 | 360.680 [13.029, 708.331] | 784.102 | 2.174 | [0.00, 3568.53] | 39.545 | 329.65 | 0.187 |
| at day 28 | 23 | 182.157 [-21.017, 385.330] | 469.838 | 2.579 | [0.00, 2269.55] | 25.79 | 229.26 | 0.045 |
| at day 56 | 23 | 201.983 [110.600, 293.400] | 211.289 | 1.046 | [0.00, 776.63] | 110.76 | 179.58 | 0.560 |
| at day 140 | 23 | 210.897 [1.579, 420.216] | 484.05 | 2.295 | [0.00, 2277.81] | 60.02 | 215.39 | 0.061 |

n: Number of non-missing values; CI: Confidence interval; SD: Standard Deviation; CV: Coefficient of Variation; Min: Minimum; Max: Maximum; IQR: Interquartile Range. Wilcoxon matched-pairs signed-rank tests on equality of pre- and post-boosting measurements.

### **Supplementary Table 35**: CXCL9 release after PBMC re-stimulation - Absolute values

| **CXCL9 (pg/mL)** | | | | | | | | |
|---|---|---|---|---|---|---|---|---|
| | n | Mean [95% CI] | SD | CV | [Min, Max] | Median | IQR | p |
| at baseline | 23 | 4061 [1981, 6140] | 4808.402 | 1.184 | [45.41, 22141.35] | 3140.14 | 3678.63 | |
| at day 7 | 23 | 7855 [5543, 10166] | 5345.966 | 0.681 | [0.00, 21261.87] | 6860.46 | 6715.72 | 0.001 |
| at day 14 | 22 | 7464 [5022, 9906] | 5506.915 | 0.738 | [100.74, 17728.13] | 6121.14 | 8854.12 | 0.010 |
| at day 28 | 23 | 6369 [4108, 8629] | 5227.419 | 0.821 | [1504.12, 18617.07] | 3824.41 | 6788.69 | 0.045 |
| at day 56 | 23 | 5177 [3246, 7109] | 4466.540 | 0.863 | [74.95, 14112.07] | 3964.49 | 7660.53 | 0.360 |
| at day 140 | 23 | 6133 [3535, 8732] | 6008.368 | 0.980 | [82.01, 24257.06] | 4211.78 | 9200.77 | 0.038 |

n: Number of non-missing values; CI: Confidence interval; SD: Standard Deviation; CV: Coefficient of Variation; Min: Minimum; Max: Maximum; IQR: Interquartile Range. Wilcoxon matched-pairs signed-rank tests on equality of pre- and post-boosting measurements.

### **Supplementary Table 36**: CXCL10 release after PBMC re-stimulation - Absolute values

| **CXCL10 (pg/mL)** | | | | | | | | |
|---|---|---|---|---|---|---|---|---|
| | n | Mean [95% CI] | SD | CV | [Min, Max] | Median | IQR | p |
| at baseline | 23 | 14684 [1462, 27906] | 30575.499 | 2.082 | [0.00, 148371.10] | 5606.22 | 16839.15 | |
| at day 7 | 23 | 40687 [24209, 57165] | 38104.866 | 0.937 | [0.00, 159910.00] | 39032.95 | 39294.19 | 0.001 |
| at day 14 | 22 | 30022 [17034, 43011] | 29294.901 | 0.976 | [388.67, 112601.60] | 20145.85 | 31668.78 | 0.003 |
| at day 28 | 23 | 23189 [12883, 33494] | 23831.276 | 1.028 | [0.00, 99593.38] | 12471.94 | 33023.07 | 0.070 |
| at day 56 | 23 | 21895 [7741, 36050] | 32732.271 | 1.495 | [392.34, 128529.90] | 7987.29 | 16906.88 | 0.410 |
| at day 140 | 23 | 31608 [15718, 47497] | 36745.028 | 1.163 | [226.39, 161925.50] | 21588.24 | 46225.42 | 0.020 |

n: Number of non-missing values; CI: Confidence interval; SD: Standard Deviation; CV: Coefficient of Variation; Min: Minimum; Max: Maximum; IQR: Interquartile Range. Wilcoxon matched-pairs signed-rank tests on equality of pre- and post-boosting measurements.

### **Descriptive statistics for Figure S3:**

**Supplementary Table 37**: IL-2 release after BAL cell re-stimulation - Absolute values

| IL-2 (pg/mL) | | | | | | | | |
|---|---|---|---|---|---|---|---|---|
| | n | Mean [95% CI] | SD | CV | [Min, Max] | Median | IQR | p |
| at baseline | 20 | 20.19 [7.19, 33.19] | 27.78 | 1.376 | [0.00, 115.88] | 17.42 | 27.36 | |
| at day 14 | 21 | 567 [-33, 1167] | 1318.00 | 2.324 | [0.00, 6190.50] | 188.71 | 463.02 | <0.001 |

n: Number of non-missing values; CI: Confidence interval; SD: Standard Deviation; CV: Coefficient of Variation; Min: Minimum; Max: Maximum; IQR: Interquartile Range. Wilcoxon matched-pairs signed-rank tests on equality of pre- and post-boosting measurements.

**Supplementary Table 38**: IL-10 release after BAL cell re-stimulation - Absolute values

| IL-10 (pg/mL) | | | | | | | | |
|---|---|---|---|---|---|---|---|---|
| | n | Mean [95% CI] | SD | CV | [Min, Max] | Median | IQR | p |
| at baseline | 20 | 6.85 [-0.37, 6.05] | 6.85 | 2.412 | [0.00, 23.54] | 0.00 | 1.25 | |
| at day 14 | 21 | 52.22 [18.66, 66.20] | 52.22 | 1.231 | [0.00, 204.06] | 26.16 | 53.40 | <0.001 |

n: Number of non-missing values; CI: Confidence interval; SD: Standard Deviation; CV: Coefficient of Variation; Min: Minimum; Max: Maximum; IQR: Interquartile Range. Wilcoxon matched-pairs signed-rank tests on equality of pre- and post-boosting measurements.

**Supplementary Table 39**: IL-12p70 release after BAL cell re-stimulation - Absolute values

| IL-12p70 (pg/mL) | | | | | | | | |
|---|---|---|---|---|---|---|---|---|
| | n | Mean [95% CI] | SD | CV | [Min, Max] | Median | IQR | p |
| at baseline | 20 | 0.84 [0.41, 1.26] | 0.91 | 1.088 | [0.00, 2.60] | 0.53 | 1.77 | |
| at day 14 | 21 | 3.85 [2.28, 5.43] | 3.46 | 0.899 | [0.00, 14.89] | 3.26 | 3.89 | <0.001 |

n: Number of non-missing values; CI: Confidence interval; SD: Standard Deviation; CV: Coefficient of Variation; Min: Minimum; Max: Maximum; IQR: Interquartile Range. Wilcoxon matched-pairs signed-rank tests on equality of pre- and post-boosting measurements.

**Supplementary Table 40**: TNF-alpha release after BAL cell re-stimulation - Absolute values

| TNF-alpha (pg/mL) | | | | | | | | |
|---|---|---|---|---|---|---|---|---|
| | n | Mean [95% CI] | SD | CV | [Min, Max] | Median | IQR | p |
| at baseline | 20 | 118098 [-101000, 337196] | 468143 | 3.964 | [0, 2094400] | 47 | 4671 | |
| at day 14 | 21 | 27898 [-17090, 72886] | 98833 | 3.543 | [0, 456203] | 624 | 9893 | 0.330 |

n: Number of non-missing values; CI: Confidence interval; SD: Standard Deviation; CV: Coefficient of Variation; Min: Minimum; Max: Maximum; IQR: Interquartile Range. Wilcoxon matched-pairs signed-rank tests on equality of pre- and post-boosting measurements.

**Supplementary Table 41**: CXCL9 release after BAL cell re-stimulation - Absolute values

| **CXCL9 (pg/mL)** | | | | | | | | |
|---|---|---|---|---|---|---|---|---|
| | n | Mean [95% CI] | SD | CV | [Min, Max] | Median | IQR | p |
| at baseline | 20 | 699 [259, 1139] | 940 | 1.345 | [0, 3742] | 214 | 1305 | |
| at day 14 | 21 | 7674 [4765, 10583] | 6391 | 0.833 | [0, 25408] | 5333 | 9780 | <0.001 |

n: Number of non-missing values; CI: Confidence interval; SD: Standard Deviation; CV: Coefficient of Variation; Min: Minimum; Max: Maximum; IQR: Interquartile Range. Wilcoxon matched-pairs signed-rank tests on equality of pre- and post-boosting measurements.

**Supplementary Table 42**: CXCL10 release after BAL cell re-stimulation - Absolute values

| **CXCL10 (pg/mL)** | | | | | | | | |
|---|---|---|---|---|---|---|---|---|
| | n | Mean [95% CI] | SD | CV | [Min, Max] | Median | IQR | p |
| at baseline | 20 | 12532 [5008, 20056] | 16076 | 1.283 | [0, 61910] | 4335 | 21731 | |
| at day 14 | 21 | 65913 [45705, 86120] | 44393 | 0.674 | [0, 158145] | 65533 | 72315 | <0.001 |

n: Number of non-missing values; CI: Confidence interval; SD: Standard Deviation; CV: Coefficient of Variation; Min: Minimum; Max: Maximum; IQR: Interquartile Range. Wilcoxon matched-pairs signed-rank tests on equality of pre- and post-boosting measurements.
